# Interpreting Breakthrough Infections Given Assortative Mixing of Partially Vaccinated Populations

**DOI:** 10.64898/2026.01.22.26344544

**Authors:** Mallory J. Harris, Akash Arani, Tapan Goel, Kejia Zhang, Stephen J. Beckett, Nathan C. Lo, Jonathan Dushoff, Joshua S. Weitz

## Abstract

Declining vaccine coverage across the United States has increased the risk of outbreaks of vaccine-preventable diseases. Even when vaccines have low primary failure rates, conventional epidemic theory predicts a strongly nonlinear, positive relationship between vaccine coverage and the fraction of breakthrough infections in vaccinated individuals. These breakthrough infections may generate misconceptions that vaccines are not working and accelerate declines in confidence and coverage. Here, we set out to test predictions of conventional epidemic theory that assumes random mixing between individuals irrespective of vaccine status. In contrast to expectations from random mixing models, we find a far lower fraction of breakthrough infections in measles outbreak data from seven states in the United States. To explore this discrepancy, we evaluate an alternative, compartmental disease model that accounts for preferential mixing (‘assortativity’) between people with the same vaccination status. The model with assortativity predicts significantly lower fractions of breakthrough infections than conventional models, consistent with observations from measles outbreak data, even when accounting for potential biases in case reporting. Next, we leverage the deviation between statewide and school-level vaccine MMR coverage across kindergartens in sixteen states, finding substantial assortativity in all cases. Our model accounting for preferential mixing predicts the total number of breakthrough infections is nonlinear, peaking at intermediate coverage below vaccine-derived herd immunity. Nationally, more than 90% of counties that report MMR coverage are above the model-predicted breakthrough-maximizing coverage, suggesting that they are at risk for increasing breakthrough infections if coverage declines. Vaccination outreach and monitoring campaigns should develop proactive strategies to contextualize breakthrough infections before low levels of primary failure contributes to population-scale increases in preventable disease.

## 1 Introduction

Vaccine coverage is declining in the United States, contributing to outbreaks of vaccine-preventable diseases (Kiang et al., 2025; Dong et al., 2025b). The ongoing measles outbreak in the U.S. exceeded 2,200 confirmed cases in 2025, spreading largely but not entirely within unvaccinated individuals. Large-scale measles outbreaks are also unfolding in Canada and Mexico, with *>*5,000 and *>*6,000 confirmed cases in 2025 alone, respectively. The Pan American Health Organization declared the loss of measles-free status and the reestablishment of endemic measles transmission in the Americas on November 10, 2025 (Pan American Health Organization, 2025). Decreasing vaccine coverage increases the outbreak risk for other vaccine-preventable illnesses, including polio and diptheria (Kiang et al., 2025). Changes in vaccine coverage may also shift the relative and total prevalence of infections in vaccinated individuals (i.e., breakthrough infections associated with vaccine failure (Heininger et al., 2012)).

Understanding the impact of reduced vaccine coverage on breakthrough infections has important implications for strategic communication, epidemiological analysis, and public health preparedness. Part of what fuels vaccine skepticism among individuals is the notion that vaccines are at best ineffective and at worst dangerous (including the idea that vaccination can *increase* the risk of infection) and therefore not worth the perceived cost (Kata, 2010). Recent viral social media posts have cited the apparently high proportion of mumps and measles infections in vaccinated individuals to suggest that vaccines are ineffective (e.g., https://archive.ph/Hxd6V, https://archive.ph/bCaaj, accessed and archived December 22, 2025). While the rate of vaccine failure is often quite low, the prevalence of infections in vaccinated versus unvaccinated individuals may be quite substantial when coverage is high (Arima and Oishi, 2018; Orenstein et al., 1985; Poland, 1994). For example, at least 15% of measles cases in New Mexico in 2025 were breakthrough infections (New Mexico Department of Health, 2025).

Conventional epidemic theory posits that the fraction of breakthrough infections in an outbreak may be leveraged to infer vaccine efficacy or vaccine coverage (Orenstein et al., 1985; Althaus and Salathé, 2015; Bhatia et al., 2025). However, such inference generally assumes random mixing within the population, whereas un-vaccinated individuals may cluster within relatively isolated communities, to which outbreaks may be confined (Munday et al., 2024; Wilson et al., 2021; Newcomer et al., 2024). Realistic assortativity within populations (i.e., limited transmission-relevant interactions between vaccinated and unvaccinated individuals) may protect vaccinated individuals from infection and reduce the number of breakthrough infections. Likewise, efforts to estimate key epidemiological parameters based on breakthrough infections may be systematically biased if they fail to account for assortativity. Changes in overall vaccine coverage and spatial clustering of unvaccinated people also have important implications for the incidence of breakthrough infections and the size of epidemics (Hiraoka et al., 2022). The proliferation of anti-vaccine rumors, including across mainstream and governmental sources, is simultaneously driving an overall increase in vaccine refusal (and nonmedical vaccine exemptions from vaccine requirements), and could reduce assortativity as vaccine hesitancy expands (Dong et al., 2025a; Carpiano et al., 2023; Fattah et al., 2026). Both trends could increase the burden of breakthrough infections as larger epidemics extend into (previously) highly vaccinated communities.

Here, we assess the link between vaccination coverage, primary failure, assortativity, and breakthrough infections. To begin, we compare data on the documented fraction of breakthrough measles infections across seven states to expectations from a compartmental model assuming random mixing. The gap between theory and state-level data suggests random mixing models are limited in their predictive power for breakthrough incidence. As an alternative, we develop and analyze an expanded epidemic model with preferential mixing. Accounting for assortativity can reconcile observations of low incidence of breakthrough infections with high coverage rates – reinforced by inference of moderate, realized assortativity from school-level MMR coverage for kindergarteners across sixteen states. While assortativity reduces the number of breakthrough infections, we find that decreases in coverage typically increases breakthrough infections, potential reinforcing anti-vaccine perspectives and catalyzing a synergistic feedback loop. Our findings support the need for principled approaches to anticipate and contextualize breakthrough infections to avoid mis-perceptions that can worsen outbreaks.

## 2. Results

### 2.1 Model-predicted vaccine coverage exceeds surveyed coverage

To understand the expected relationship between breakthrough infections and vaccine coverage, we assess a conventional epidemic model of disease transmission and vaccination for a vaccine with primary failure rate *ϵ* and population-wide vaccine coverage *p* (see Equation S1 for equations and Figure S1 for a compartmental diagram). We consider the relative and total number of breakthrough infections at equilibrium, focusing on parameters consistent with measles and the MMR vaccine (Table S1, with initial conditions in Table S2).

In instances where vaccine coverage is insufficient to eliminate a disease, it is possible to link the fraction of breakthrough infections (relative to total infections), *f*_*V*_, to vaccine coverage, *p*. Assuming no assortativity, the fraction of breakthrough infections increases with vaccine coverage, potentially sharply, such that 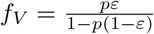 (Figure 1A, subsection S1.2). For example, if coverage drops to *p* = 0.9 assuming that the MMR vaccine has failure rate *ε* = 0.03 (McLean et al., 2013) then *f*_*V*_ ≈ 0.21 of total infections should be breakthrough infections. The positive relationship between coverage and the breakthrough fraction is maintained when two doses of a vaccine are administered (consistent with MMR), and the fraction of breakthrough cases that only received one dose is consistently greater than the fraction of two-dose breakthroughs (section S2).

**Figure 1:**
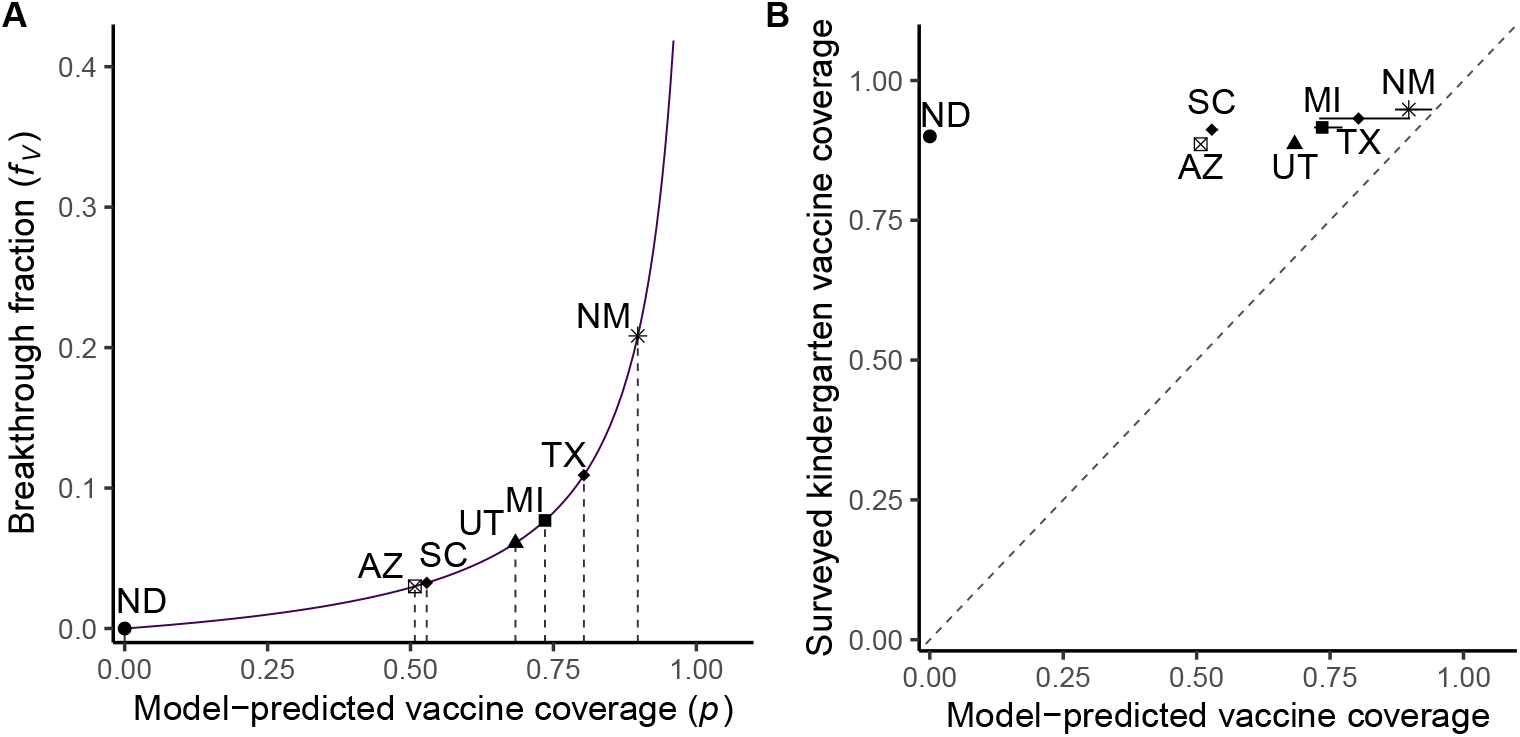
The model-predicted vaccine coverage is consistently below the surveyed kindergarten vaccine coverage. (A) The predicted positive relationship between vaccine coverage (*p*) and the breakthrough fraction (*f*_*V*_). For each of the seven states included in the analysis, the point indicates the reported breakthrough fraction for measles in 2025 and the vertical line corresponds to the model-predicted vaccine coverage. Model predictions are for parameters consistent with measles and the MMR vaccine: 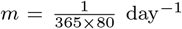, *ε* = 0.03, *γ* = 0.1 day^−1^, *β* = 15× (*γ* + *m*) day^−1^ (i.e., ℛ_0_= 15). The population has size 10M and there is no assortativity (*ϕ* = 0). See Figure S4 for a supplemental analysis using different values of ℛ_0_ and *ϵ*. (B) Comparison of model-predicted vaccine coverage to surveyed kindergarten MMR coverage for school year 2024-2025. The point indicates the breakthrough fraction across all cases with known vaccination status and the horizontal bars correspond to uncertainty in the breakthrough fraction given that some cases have unknown vaccination status (see section 4 for further explanation). The dashed line indicates perfect correspondence.

We revisit the interplay between breakthrough infections and changes in coverage in the context of the measles outbreaks in the United States in 2025. Prior work assuming random mixing has leveraged realized measurements of the fraction of breakthrough infections to infer failure rates (Orenstein et al., 1985) and vaccine coverage (Althaus and Salathé, 2015). The reported fraction of breakthrough infections across seven states ranges from 0 – 0.21. As a result, the theoretically inferred vaccine coverage rate, *p*, assuming random mixing falls below 80% in five states and below 60% in three states (Figure 1A). In contrast, the surveyed kindergarten vaccine coverage, *p*_*school*_, exceeded 85% in all states (Figure 1B). Theoretical predictions for vaccine coverage are substantially below surveyed measures for all states, with the exception of New Mexico.

### 2.2 State-level breakthrough fraction and vaccine coverage are consistent with moderate assortativity

We hypothesize that non-random mixing based on vaccination status is one possible explanation that may help resolve the discrepancy between model-predicted and surveyed vaccine coverage. We set out to test this hypothesis by incorporating assortativity into an expanded model of disease dynamics in a partially vaccinated population. In practice, we subdivide the population into vaccinated and unvaccinated groups and incorporate a preferential mixing rate *ϕ*, which corresponds to the share of an individual’s contacts that will be reserved for people with concordant vaccination status (versus contacts with the entire population; Figure S1). Using the same disease parameters in Figure 1 for measles, we find that assortative mixing reduces the expected fraction of breakthrough infections for all values of coverage (Figure 2). These results were robust to alternative assumptions on ℛ_0_ and primary failure rate (vaccine efficacy) (Figure S4). Across states, the relationship between surveyed vaccine coverage and the breakthrough fraction is positive. As hypothesized, observed low breakthrough fractions (below 0.2) at high vaccine coverage rates (above 85%) are consistent with moderate to high levels of assortativity in mixing relevant to measles transmission (*ϕ >* 0.6) (Figure 2).

**Figure 2:**
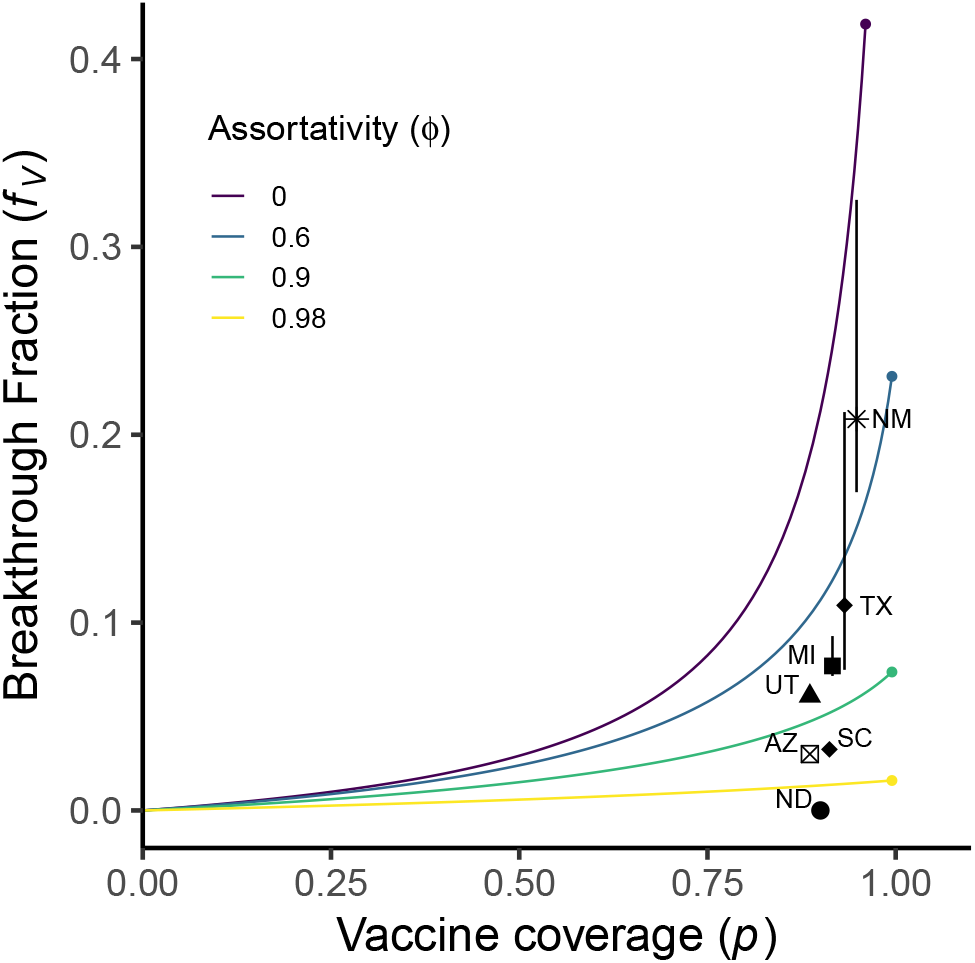
Assortativity reduces the breakthrough fraction. Lines illustrate the relationship between vaccine coverage (*p*) and the breakthrough fraction (*f*_*V*_) for different levels of assortativity (*ϕ*), ranging from 0 (no assortativity) to 0.98 (high assortativity). Parameters are consistent with measles and the MMR vaccine (see Table S1). See Figure S3 for a supplemental analysis of the effects of disparate case detection based on vaccination status and see Figure S4 for a supplemental analysis using different values of ℛ_0_ and *ϵ*. Black points indicate surveyed kindergarten vaccine coverage for school year 2024-2025 against the reported breakthrough fraction for measles in 2025 and vertical bars correspond to uncertainty in the breakthrough fraction given that some cases have unknown vaccination status (see Methods and Figure 1 for more information).

The observed breakthrough fraction will tend to be lower if breakthrough infections are more likely to be underreported compared to infections in unvaccinated individuals (Figure S3). However, even if breakthrough cases are twice as likely to be unreported (compared to unvaccinated cases), the relationship between vaccine coverage and breakthrough fraction would be consistent with moderate to high assortativity (*ϕ >* 0.5) across most states (except for Texas and New Mexico). These findings suggest the need to estimate transmission-relevant assortativity within states where measles outbreaks have occurred.

### 2.3 Differences between state- and school-level vaccine coverage are consistent with moderate assortativity

We estimated realized levels of assortativity relevant to measles transmission in the U.S., by analyzing vaccine coverage data for kindergarteners across sixteen states (including four of the seven shown in Figure 1 and Figure 2) over a multiyear period, (see Sec. 4). We estimated assortativity for each state annually by comparing school-level and statewide kindergarten MMR coverage and taking a population-weighted mean across school-level values (Equation 2). School-based assortativity estimates 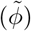 exceeded 0.2 in all cases, indicating moderate assortativity (Figure 3). The average, estimated assortativity since the 2019 – 2020 school year ranged from 0.27 in Michigan to 0.60 in Maryland (median: 0.37; mean: 0.39) (Figure 3, Table S3). States with greater estimated assortativity also had a lower breakthrough fraction, as expected (compare Figure 2 and Figure 3; ordering of states by ascending estimated assortativity or descending breakthrough fraction is: Michigan, Utah, South Carolina, and North Dakota). Estimated assortativity values are significantly positive in all cases. However, estimated assortativity was consistently below model-predicted values as inferred from the fraction of breakthrough infections (0.6 or greater, see Figure 2) – an issue we return to in the Discussion. We did not find evidence in any state for significant changes in estimated assortativity over time (Table S3, Figure S6).

**Figure 3:**
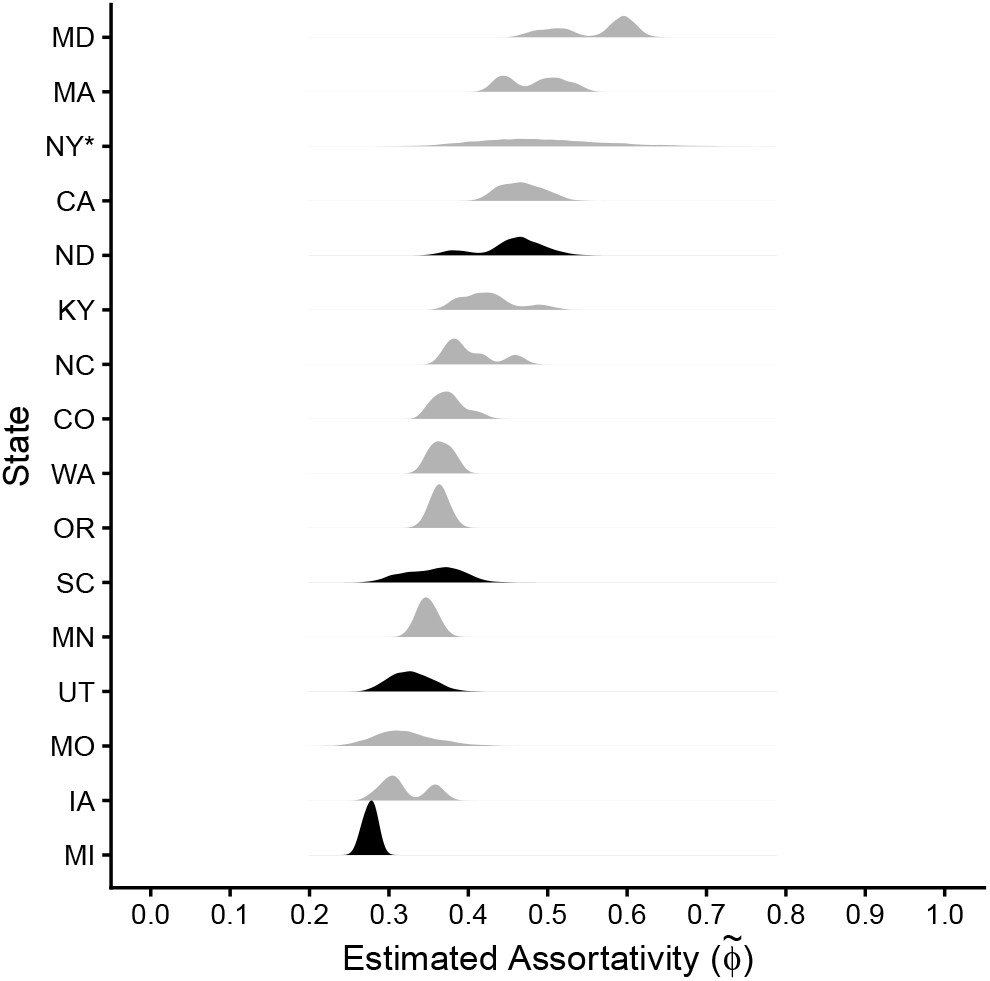
Estimated assortativity across sixteen states is moderate. Distribution of boostrapped estimated assortativity values 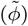 across all years available for each state, arranged vertically by descending estimated assortativity. Values of 0 would correspond to no assortativity (i.e., all schools have the same vaccine coverage) while values of 1 would correspond to complete separation between vaccinated and unvaccinated populations. The four states in black (ND, MI, SC, and UT) also had measles epidemics in 2025 and were included in the analysis of breakthrough fractions (see Figure 1 and Figure 2). *New York excludes New York City. See Table S5 for information on data sources and limitations.

### 2.4 Breakthrough infections are maximized at intermediate vaccine coverage and low assortativity

We leverage both empirical and model findings to explore scenarios of decreasing coverage and assortativity on the total number of breakthrough infections. As a baseline assuming random mixing, total infections (regardless of vaccination status) and infections in the unvaccinated population increase monotonically with decreasing coverage below the herd immunity threshold 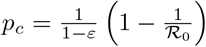 (Anderson and May (1991), subsection S1.3). As a result, the total number of breakthrough infections should follow a unimodal function of coverage and peak at a level 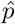 below the vaccine-induced herd immunity threshold, 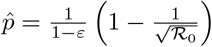 (Figure 4 and subsection S1.3). This relationship remains unimodal in the presence of assortativity. As shown in Figure 4, assortativity typically decreases the total number of breakthrough infections, although it can also make elimination more difficult when vaccine coverage is high (see Figure S7). Assortativity also generally decreases the vaccine coverage level 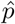 at which breakthrough infections are maximized and the peak number of breakthrough infections (although 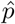 is nearly constant between 0.2 and 0.6, the range of assortativity values estimated across school-level data). However, 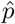 may be greater when there is some assortativity compared to no assortativity. The unimodal relationship between the number of breakthrough infections and vaccine coverage holds across different ℛ_0_ and vaccine failure rates (Figure S5).

**Figure 4:**
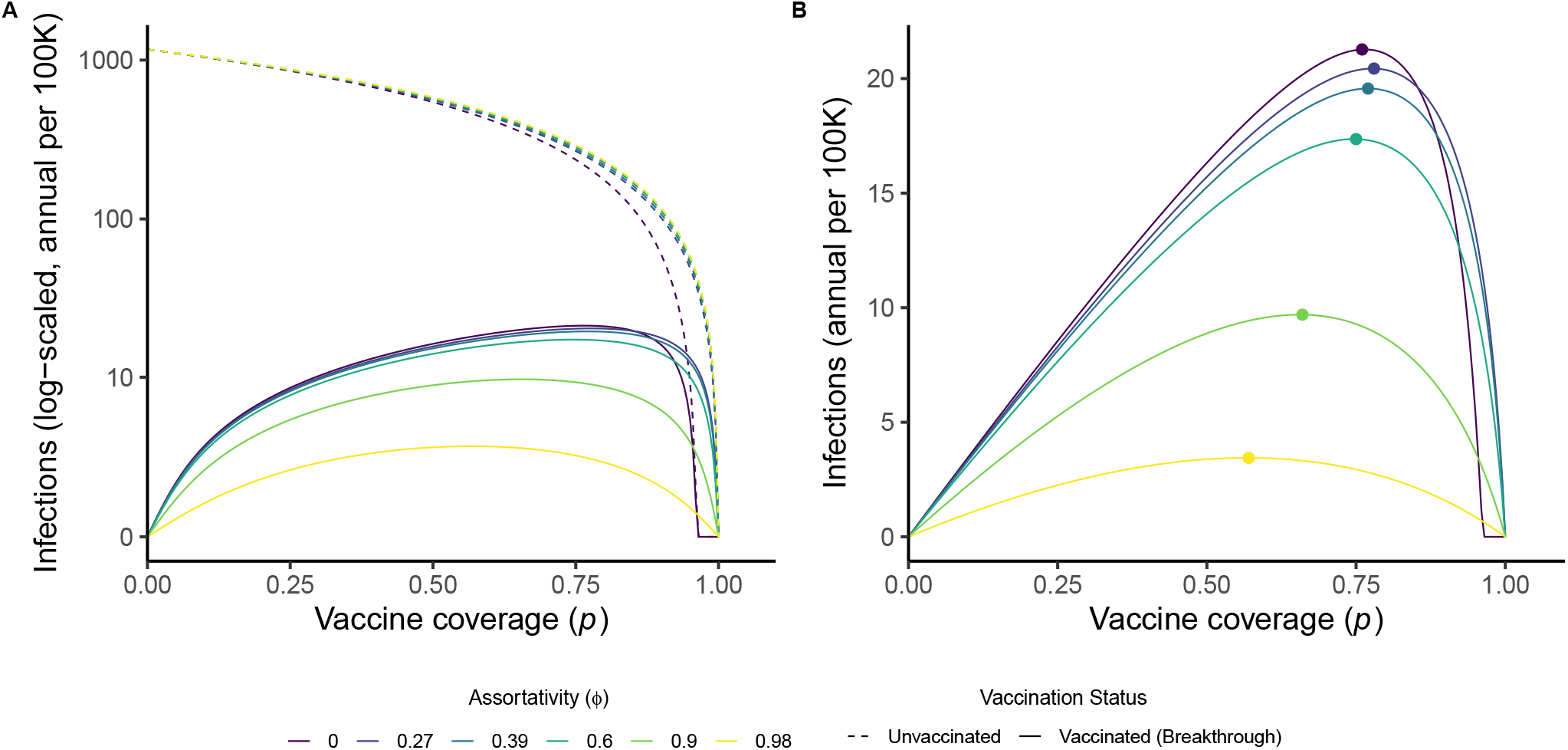
Breakthrough infections peak at intermediate vaccine coverage. Lines indicate annual new cases per 100K people depending on vaccine coverage (*p*) and assortativity (*ϕ*, colors). The left panel shows log-scaled cases in unvaccinated people (dashed lines) and vaccinated people (solid lines, breakthrough infections). The right panel shows linear-scaled breakthrough infections, with points corresponding to the peak number of breakthrough infections for different levels of assortativity 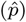. Parameters are consistent with measles and the MMR vaccine (ℛ_0_ = 15 and failure rate *ϵ* = 0.03, see Table S1). See Figure S5 for a supplemental analysis using different values of ℛ_0_ and *ϵ*.

We predict that decreasing vaccine coverage could lead to increases in the *total* number of breakthrough infections even as the *relative* fraction of breakthrough infections decreases. In the case of measles, assuming ℛ_0_ = 15 and *ε* = 0.03 and no assortativity, *p*_*c*_ ≈ 0.96 and 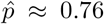, i.e., breakthrough infections should increase rapidly as coverage decreases moderately (i.e., by 5%-15%). Moreover, if coverage drops such that 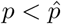 then public health efforts to increase coverage are predicted to lead to *more* breakthrough infections given the unimodal relationship between coverage and the total number of breakthrough infections. This response arises as a result of the balance between the protective benefits of increasing vaccinations that decrease the total burden of illness while increasing the size of the vaccinated population.

At assortativity consistent with the mean estimated value across states (Figure 3, 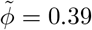), the critical coverage level *p*_*c*_ exceeds 0.99 (*p*_*c*_ = 0.9993) and the breakthrough infection-maximizing coverage 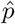 is approximately 0.77. Of the 2,141 counties in the United States that reported MMR coverage in kindergarteners for school year 2022-2023, fewer than 3% of counties (55 counties) had coverage exceeding assortativity-adjusted *p*_*c*_, while nearly 92% of counties (1,964 counties) have coverage between assortativity-adjusted *p*_*c*_ and 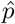 (Figure 5). Together this means that over 94% of counties have coverage exceeding 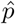 (2,019 counties for the value of 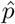 derived using the national mean estimated *ϕ*; with a range between 2,009 to 2,041 counties for values when accounting for variation in assortativity across states, see Table S3). These counties are at risk of increased breakthrough infections if vaccine coverage declines further (Figure 5).

**Figure 5:**
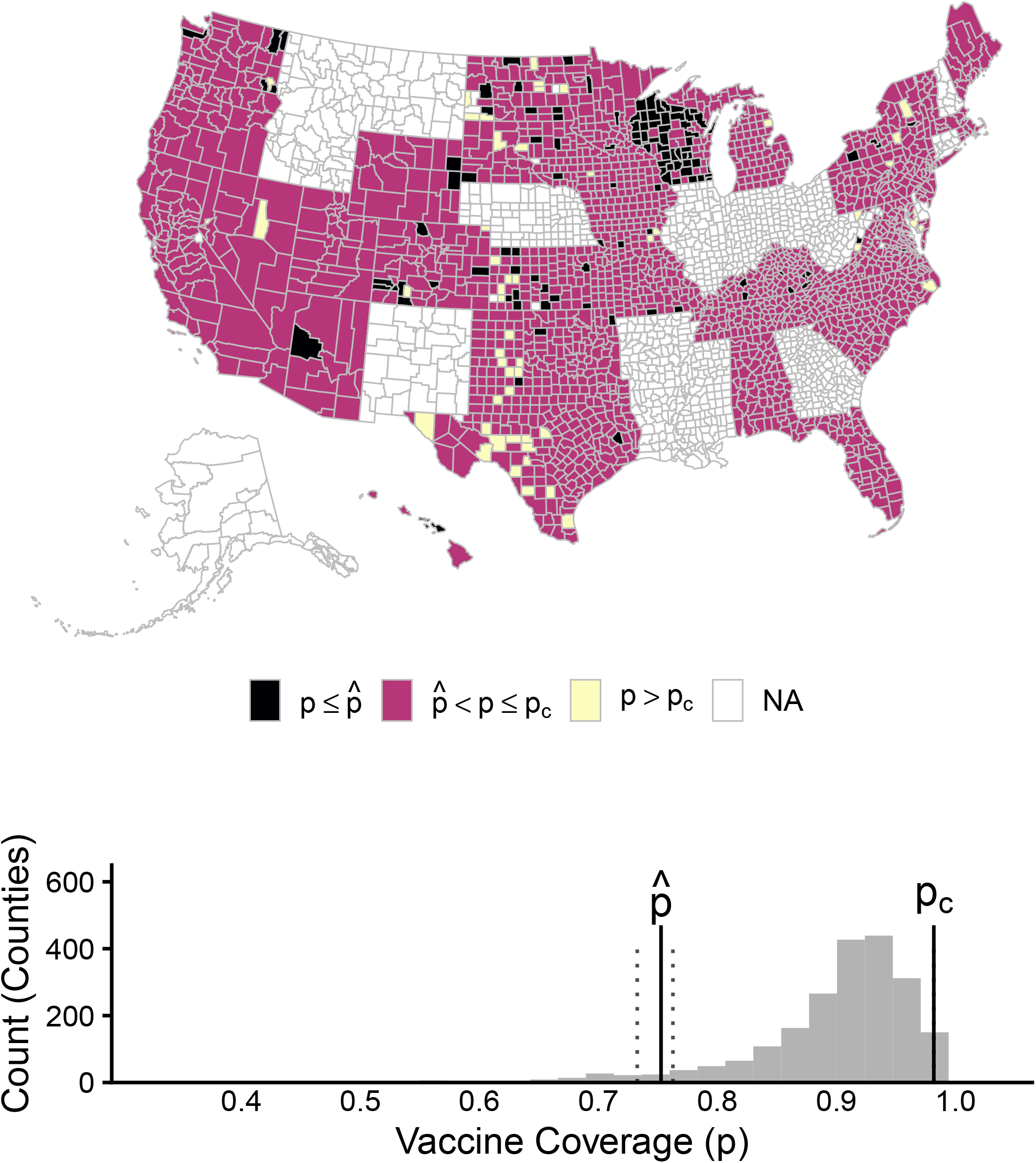
Most counties have vaccine coverage above 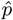, meaning more breakthrough infections could occur if coverage declines. In the map, counties are shaded depending on how their kindergarten MMR coverage for school year 2022-2023 (Dong et al., 2025b) to the critical coverage 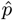 and the vaccine-induced herd immunity threshold *p*_*c*_ calculated for the mean assortativity estimated across states 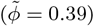 and realistic parameters for measles and the MMR vaccine (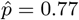 and *p*_*c*_ *>* 0.99). Counties without available data for this time period (across 13 states) are white. The bottom figure is a histogram of vaccine coverage across the 2,141 counties that reported vaccine coverage and the vertical lines indicate 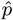 and *p*_*c*_. The dotted lines indicate the range of values for 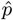 for when *ϕ* is between 0.27 and 0.60 (as estimated from school-level data, see Figure 3). Across the range of estimated *ϕ* values, *p*_*c*_ is nearly invariant.

## 3 Discussion

Declining vaccine coverage and expanded vaccine hesitancy could lead to more breakthrough infections and increase the risk of a positive feedback loop driving vaccine hesitancy and increases in vaccine-preventable disease. Analysis of an epidemic transmission model with preferential mixing within groups of vaccinated and unvaccinated individuals shows that once coverage has dropped below vaccine-induced herd immunity thresholds, decreasing vaccination rates can increase breakthrough infections, undermining public health objectives. We also find evidence of preferential mixing (i.e., ‘assortativity’) based on vaccination status – supported by comparing data on recent outbreaks to transmission model predictions and by analyzing differences between state- and school-level vaccination coverage. Vaccine assortativity makes outbreaks like the current U.S. measles outbreak more likely, but also reduces the proportion of breakthrough infections. Moving forward, reduced vaccine coverage and decreases in assortativity could lead to substantial increases in the total number of breakthrough infections. The present analysis comes with limitations. We model primary failure as an all-or-nothing process (where people are either fully protected or completely unprotected), but disease dynamics are less sensitive to the mode of failure when vaccine effectiveness is relatively high, as is the case for MMR (Lee et al., 2025). We do not consider secondary failure (i.e., waning immunity), which may be particularly important in highly vaccinated populations where incidence has been relatively low and cases occur predominantly in adults (Paunio et al., 2003; Yang et al., 2020; Leung et al., 2025; Robert et al., 2024). Nonetheless, the results are generally robust to the primary failure rate and ℛ_0_, suggesting the relationships between vaccine coverage, assortativity, and breakthrough infections are relevant across varied contexts (Figure S4, Figure S5). Following the studies used to estimate vaccine failure rates, we focus on detectable breakthrough cases, which are typically symptomatic (McLean et al., 2013; Uzicanin and Zimmerman, 2011; De Serres et al., 1995; Yeung et al., 2005; Vitek et al., 1999; De Serres et al., 2012; Sutcliffe and Rea, 1996). Although subclinical measles infection have been observed in vaccinated individuals (Pedersen et al., 1989), they are unlikely to contribute substantially to onward transmission (Lievano et al., 2004). Efforts to characterize the precise mechanism of vaccine failure and implications for breakthrough infections are especially important in the context of discussions about next-generation vaccine development and updated measles vaccination strategies in elimination contexts (Paunio et al., 2003; Yang et al., 2020; Poland and Jacobson, 2012). Likewise, community surveys may be necessary to quantify the extent to which measles infections are reported for unvaccinated and vaccinated individuals.

The epidemic transmission model may also underestimate the benefits of current vaccination practices and the risks of assortativity based on vaccine status. Although the epidemic model assumes a single dose vaccination for simplicity, MMR, and most other childhood vaccines, require multiple doses. In an expanded two-dose model, we show that a larger share of breakthrough infections occur in people who have only received a single dose, underscoring the importance of counting breakthrough infections based on dose number in order to avoid underestimating the effectiveness of complete vaccination (Figure S2). We assume that breakthrough infections are identical to infections in unvaccinated people. In practice, breakthrough infections may be less infectious and associated with fewer symptoms (Cherry and Zahn, 2018; Leung et al., 2025; Sundell et al., 2019; Evans et al., 2024), while increased activity of vaccinated people who may have more mild symptoms or assume that they are unable to become infected could lead to higher effective transmission rates (Park et al., 2020; Pedroza-Meza et al., 2026). Also, children below vaccination age constitute a sizable proportion of the unvaccinated population and may be at greater risk of severe illness, meaning that cases in unvaccinated individuals are especially concerning (Leung et al., 2025).

Considering a variety of epidemiological outcomes may provide additional insight into the impacts of vaccination campaigns depending on various goals, which may depend on the disease or vaccine (e.g., reducing clinical severity or preventing infections in certain groups). Although we focus on the potential for diminished assortativity to shift disease burden toward vaccinated people in the context of sustained transmission, our model does not consider epidemic risk following re-introduction, which may be considerably greater in spatially clustered populations (Truelove et al., 2019; Chen and Bento, 2026). This work underscores the importance of considering the combined impacts of vaccine coverage and assortative mixing on disease risk both across the population at large and among vaccinated people (Hiraoka et al., 2022).

While this study suggests that assortativity may partially explain the tendency for the model-predicted fraction of breakthrough infections to exceed observed values based on state-level coverage, limitations in data collection may also contribute to the gap between model-predicted and estimated assortativity (see Figure 3). Although measles cases in the United States in 2025 exceeded cases over the past two decades and these outbreaks remain concerning from a public health standpoint, the small number of cases reported in any given state further complicate attempts to draw inference based on the breakthrough fraction. Underdetection may also bias estimates; specifically, relatively greater underreporting of breakthrough infections relative to infections in unvaccinated individuals (for example, because breakthrough infections may be more mild) could additionally suppress the observed breakthrough fraction, while reluctance to report infections in unvaccinated individuals could bias estimates in the other direction (Figure S3). This analysis suggests that breakthrough infections are underdetected relative to infections in vaccinated individuals across most of the states examined (Figure S3). State-level reports of vaccine coverage may exceed coverage within local communities where outbreaks occurred (Fitzpatrick et al., 2025). For example, Mohave County, where the outbreak in Arizona has been centered, had county-level MMR coverage in kindergarteners of 76% compared to 89% coverage at the state level (Dong et al., 2025b; Centers for Disease Control and Prevention, 2025b). Although vaccine coverage is generally included in outbreak reports (Rahimi et al., 2025) and school-level coverage for kindergarteners is reported by several states, we recommend improvement of assessment and reporting of local vaccine coverage (including in school-aged children of different grades) and standardization of reporting practices across regions (Dolan et al., 2019).

Efforts to measure vaccine coverage in preschool and adult populations and describe coverage and contact patterns outside of school settings (including contact rates dependent on vaccination status) are necessary to improve understanding of transmission dynamics, and underlying risk (Gastañaduy et al., 2020; Salmon et al., 2006; Bednarczyk and Sundaram, 2025; Zhou et al., 2026; Hiraoka et al., 2022). Of particular relevance, the vaccine series commences at twelve months of age and is completed between ages four and five (Centers for Disease Control and Prevention, 2025a), meaning that contact rates by age, binned by vaccine eligibility, are especially important to understand the potential for transmission from contact with unvaccinated individuals (Taube et al., 2025; Nelson et al., 2021; Andrejko et al., 2022). Epidemiological analyses at granular geographic scales (i.e., sub-state and even sub-county) will help guide response, given that fine-scale heterogeneity in vaccine coverage exists and substantially shapes epidemic dynamics (Fitzpatrick et al., 2025; Masters et al., 2020; Truelove et al., 2019; Fattah et al., 2026; Dong et al., 2025b; Zhou et al., 2026; Chen and Bento, 2026).

The combination of theory and data-driven analyses suggest that projected increases in the number of break-through infections could necessitate shifts in surveillance and quarantine strategies that currently assume infections will be rare in vaccinated people (Cherry and Zahn, 2018; Cassini et al., 2024; Poland and Jacobson, 2012; Leung et al., 2025). Research to characterize how information about breakthrough infection affects vaccine decision-making, coupled with models that incorporate the behavior-disease feedback loop between breakthrough infections, vaccine hesitancy, and declining coverage, may help inform interventions (Pedroza-Meza et al., 2026). Efforts to increase public trust in vaccination and address the specific concerns of different communities are especially urgent to counter the diffusion of vaccine hesitancy (which could also decrease assortativity) (Carpiano et al., 2023; Hijano et al., 2025). States may also improve vaccine uptake by implementing evidence-based practices including elimination of nonmedical vaccine exemptions, expanded authorization of vaccinators, and investment in expanded outreach campaigns to reach undervaccinated communities (Cunniff et al., 2023). We recommend that communication and monitoring efforts anticipate the link between changes in coverage and increases in breakthrough incidence. Improving baseline expectations and managing interpretations of break-through infections may help avoid avoidable errors in vaccination initiatives.

## 4 Methods

### 4.1 Epidemic transmission model with assortativity

We propose an epidemic transmission model that partitions individuals into the following compartments: susceptible and unvaccinated (*S*_*U*_), infected and unvaccinated (*I*_*U*_), susceptible and vaccinated (*S*_*V*_), infected and vaccinated (*I*_*V*_), recovered from an infection without vaccination (*R*_*U*_), and vaccinated or recovered from a breakthrough infection (*R*_*V*_). We assume that infection confers sterilizing immunity. We track the size of the population of individuals who are unvaccinated (*N*_*U*_ = *S*_*U*_ + *I*_*U*_ + *R*_*U*_) or vaccinated (*N*_*V*_ = *S*_*V*_ + *I*_*V*_ + *R*_*V*_) and assume a stable population size (*N*_*T*_ = *N*_*U*_ + *N*_*V*_). Births occur at a constant rate *b*, to balance population-level deaths (independent of infection status) happening at a constant per capita rate (i.e., *b* = *mN*_*T*_). Unvaccinated people are vaccinated with probability *p* (the vaccine coverage).

We assume that the vaccine has no effect (primary failure) with a probability *ε*, and provides sterilizing immunity otherwise. Contacts may be assortative, meaning that people are more likely to contact others who share their vaccination status. Assortativity is parameterized by *ϕ*, corresponding to the proportion of contacts reserved for people with concordant vaccination status versus the entire population (Busenberg and Castillo-Chavez, 1991; Dushoff and Levin, 1995). We allow *ϕ* to vary from 0 (no asssortativity; random mixing) to 1 (full assortativity; no contact between people with discordant vaccination status). Accounting for assortativity, infected individuals contribute equally to the force of infection with transmission coefficient *β* irrespective of their vaccine status. Finally, infected individuals recover at the same per capita rate *γ*. The complete model is:

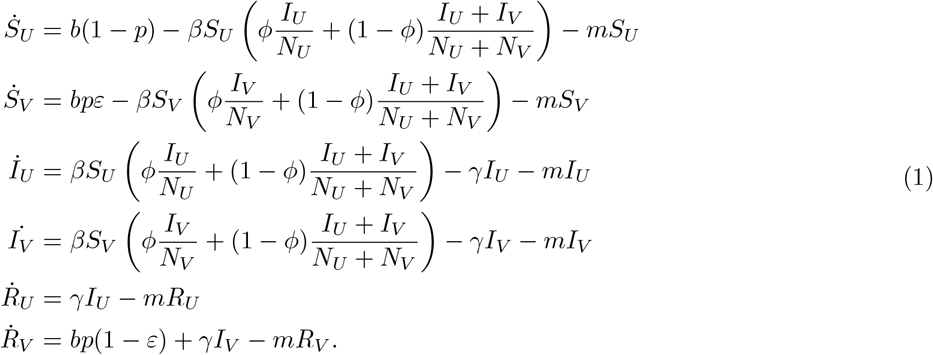

The equations were solved numerically using the ode23t solver in MATLAB (The MathWorks, Inc., 2025) for the range of parameter values and initial conditions provided in Table S1 and Table S2. The time step-size was set adaptively by the solver, under the constraint that the relative error tolerance for each integration step was set to 10^−8^ for each state variable and all state variables had to be non-negative at all times. To obtain the equilibrium system state for each set of parameter values, the differential equations were integrated up to *T* = 5 ×10^5^ days, which was adequate time for the system to reach equilibrium. The state of the system at time *T* was taken to be the equilibrium state. The MATLAB parallel computation toolbox was used to solve for steady states across different parameter values. Additional analyses and figures were conducted in RStudio using R version 4.2.3 (Posit team, 2025; R Core Team, 2021).

### State-level outbreak and vaccination data

We compiled publicly available data from twelve state departments of health that CDC lists as providing realtime updates on measles case reports (https://www.cdc.gov/measles/data-research/index.html, last accessed 15 December 2025) (see Table S4 for data and sources). Six of the states (Michigan, New Mexico, North Dakota, South Carolina, Texas, and Utah) report cases at the state level categorized by vaccination status (where vaccinated indicates at least one dose of MMR) and one state (Arizona) reports the total number of cases and the percentage of cases in unvaccinated people. All states except for Texas disaggregate unvaccinated cases from cases with unknown vaccination status. For our calculation, we assumed at least half of the unknown/unvaccinated cases were unvaccinated and classified the remainder as unvaccinated. We calculated the fraction of breakthrough infections (*f*_*V*_) as the fraction of all known cases occurring in people with at least one dose of MMR. Across all states, we estimated a lower bound on *f*_*V*_ by assuming that people with unknown status are vaccinated at one third the rate of cases with known status; we estimated an upper bound by assuming they were three times more likely to be vaccinated.

### School-level vaccination data

We examined school-level vaccine coverage to approximate assortativity in school attendance based on vaccine status. We collected data by first examining state health department websites across all fifty states and then requesting data directly if they were not available on public dashboards. Annual MMR coverage (two doses completed) and number of students at the school level for kindergartens across the state were available for twelve states. Michigan and Iowa report coverage for all required vaccines rather than just MMR, Missouri data are at the zip code level, South Carolina reports data for a sample of schools, and several states suppressed data from schools with few students enrolled for privacy (see Table S5 for detailed information about school-level data). In total, we included sixteen states in the analysis. Preferential attendance based on vaccination status (*ϕ*_*i*_) was approximated by comparing school-level vaccine coverage (*p*_*i*_) to state-level coverage (*P*, estimated from each year’s school-level data):

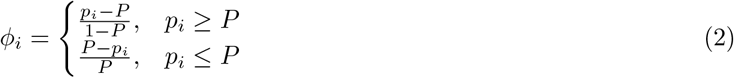

We then took a state-wide, annual average of this metric 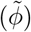, weighting by the share of the state’s children attending each school. We bootstrapped calculations by resampling 1,000 times from schools in a given state and year and taking the 95% confidence interval across this distribution of values. In a supplemental analysis, we tested whether estimated assortativity has significantly changed in any state since the 2019 – 2020 school year (with Bonferroni correction, *α* = 0.05*/*16 = 0.003).

## Data Availability

All associated code has been uploaded to Github at https://github.com/WeitzGroup/Breakthrough-Infections/ and is archived on Zenodo at https://doi.org/10.5281/zenodo.20614166. Publicly available outbreak and vaccination data across six states are included in the repository, while data obtained via correspondence with public health departments in ten states are withheld.

https://doi.org/10.5281/zenodo.20614166

## Acknowledgments

M.J.H., S.J.B., and J.S.W. are investigators at the University of Maryland-Institute for Health Computing, which is supported by funding from Montgomery County, Maryland and The University of Maryland Strategic Partnership: MPowering the State, a formal collaboration between the University of Maryland, College Park and the University of Maryland, Baltimore. This work is supported by a Simons Foundation grant to J.S.W. (MPS-SIP-00930382). N.C.L. is supported by the National Institutes of Health (NIAID) New Innovator Award (DP2AI170485).

We thank state public health departments across the country, which provided measles infection and/or vaccination data for this analysis: Arizona Department of Health Services; California Department of Public Health; Colorado Department of Public Health and Environment; State of Iowa Health and Human Services (Open Record request A26-261, 12.13.2025); Kentucky Department for Public Health Immunizations Branch; Maryland Department of Health Center for Immunization; Massachusetts Department of Public Health (data and technical guidance from Joshua Norville and Christopher Tocci from the Massachusetts Bureau of Infectious Disease and Laboratory Sciences); Michigan Department of Health and Human Services (data and technical guidance from Taylor Olsabeck, VPD Immunization Epidemiology Section Manager and Thrishika Balasubramanian, VPD Epidemiologist); Minnesota Department of Health Infectious Disease Epidemiology, Prevention and Control Division; Bureau of Data Modernization and Interoperability and the Bureau of Immunizations at the Missouri Department of Health and Senior Services; New Mexico Department of Health; New York State Department of Health; North Carolina Department of Health and Human Services Division of Public Health; North Dakota Department of Health and Human Services; Oregon Health Authority Immunization Program; South Carolina Department of Public Health Bureau of Communicable Disease Prevention and Control; Texas Department of State Health Services; Utah Department of Health and Human Services Office of Communicable Diseases and Immunization Program; and Washington State Department of Health. Analyses and conclusions are those of the authors and do not represent the views of any state department of health.

## Supplementary Information for

### S1 Mathematical supplement

#### S1.1 Breakthrough infections in endemic steady state, in the absence of assortativity

As in the main text, we investigate the burden of breakthrough infections as a function of disease properties, vaccination coverage, and vaccine failure rates by developing a compartmental model as follows (see compartmental diagram, Figure S1):

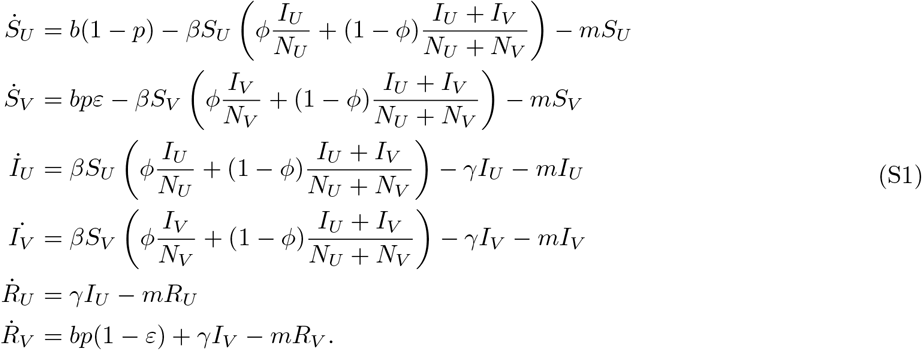

We first analyze the endemic steady state when there is no assortivity, i.e. *ϕ* = 0, to understand how vaccine coverage (*p*) and vaccine failure (*ε*) impact prevalence of breakthrough infections. If 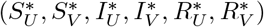 denotes the endemic equilibrium of Equation S1 in the absence of assortivity, then:

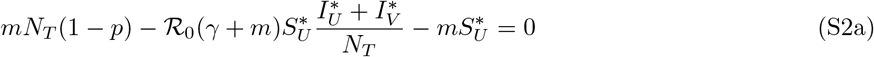

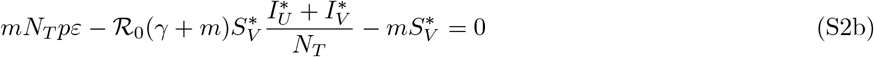

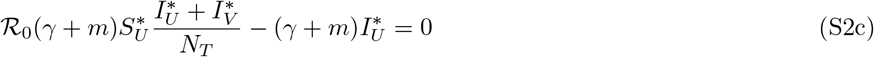

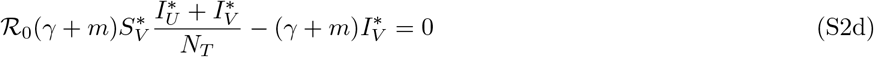

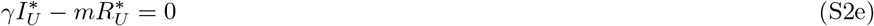

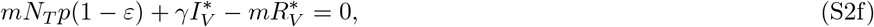

where, 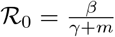 and *b* = *mN*_*T*_.

Adding Equation S2c and Equation S2d:

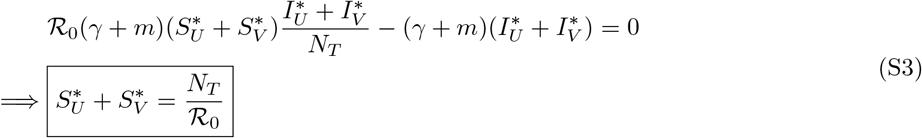

Further, adding Equation S2a and Equation S2b:

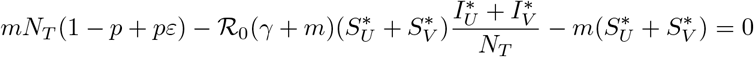

Substituting for 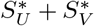 from Equation S3 and rearranging:

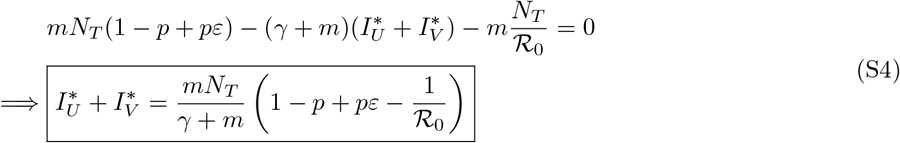

Substituting Equation S4 in Equation S2a,

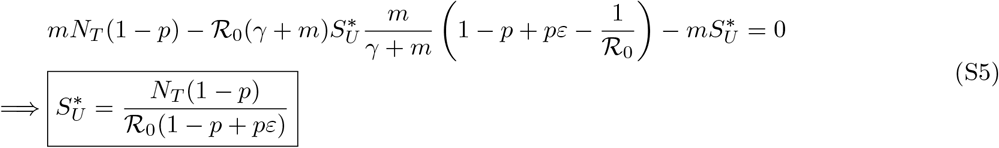

Substituting Equation S5 in Equation S3,

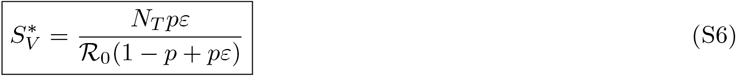

Substituting 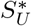 from Equation S5 and 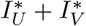 from Equation S4 into Equation S2c,

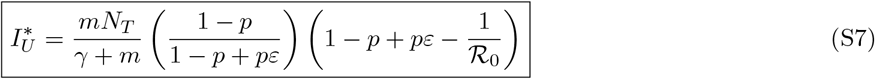

Solving for 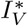 by substituting 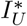 from Equation S7 in Equation S4:

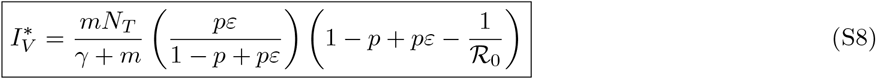

#### S1.2 Relative breakthroughs at equilibrium

The fraction of breakthrough infections at equilibrium is:

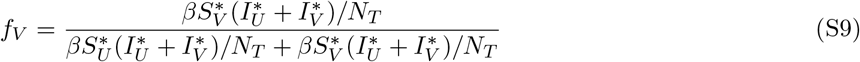

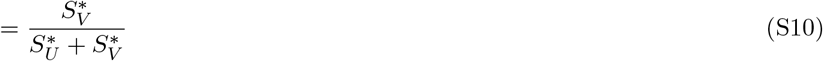

Substituting for 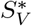 and 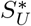 from Equation S5 and Equation S6 respectively,

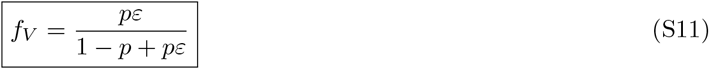

For very high vaccine failure rates, i.e., *ε* → 1, *f*_*V*_ ≈ *p*. This is to be expected – if the vaccine doesn’t work, then all individuals are equally susceptible and the proportion of infected individuals who are vaccinated would be the same as the proportion of individuals who are vaccinated (Figure S4). However, if the vaccine failure rate is low, i.e., 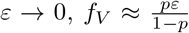, meaning that the fraction of breakthrough infections is a convex function of the coverage.

#### S1.3 Vaccine coverage required for herd immunity (*p*_*c*_)

If the vaccine coverage exceeds the herd immunity threshold (*p*_*c*_), the disease would not emerge if an infectious individual were added to the disease-free equilibrium (DFE) state. For this to happen, both eigenvalues of the infectious subsystem defined by (*I*_*U*_, *I*_*V*_) must be negative, when the system is linearized around the DFE. The equation for the infectious subsystem linearized around the DFE is:

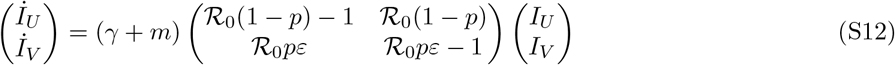

For both eigenvalues of this system to be negative,

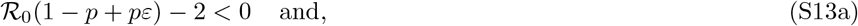

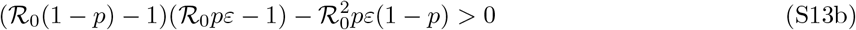

For both these conditions to hold,

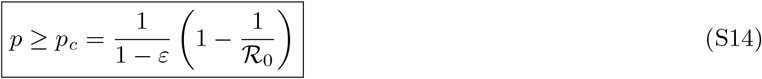

If the vaccine coverage exceeds *p*_*c*_, the disease does not become endemic. Note that *p*_*c*_ is an increasing function of *ε*. This means that as the failure rate of the vaccine increases, a higher level of vaccine coverage is required to prevent the disease from becoming endemic (Figure S5).

#### S1.4 Coverage at peak breakthrough infections (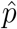)

We consider the degree of coverage, 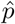, that corresponds to the highest level of breakthrough infections. At 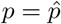, then 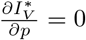. Plugging in 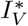 from Equation S8:

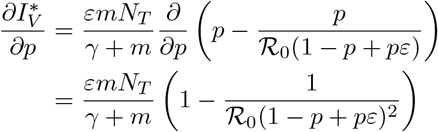

Substituting 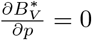 and 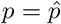:

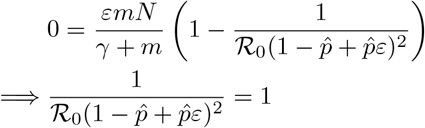

Rearranging the terms,

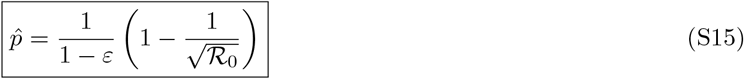

If ℛ_0_ *>* 1, 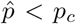. This means that the number of breakthrough infections peaks at vaccination coverage below the threshold for herd immunity, as shown in Figure 4.

### S2 Supplemental analysis: Two-dose post-maternal immunity framework

We propose a two-dose epidemic transmission model to characterize early-life infection dynamics following the waning of maternal antibodies and the sequential administration of a two-dose childhood vaccine (e.g., MMR) (LeBaron et al., 2007). The complete system of equations is given by:

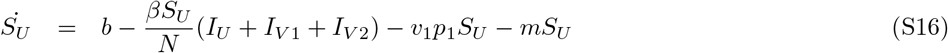

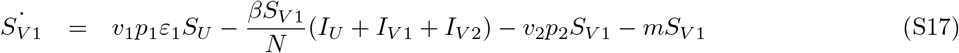

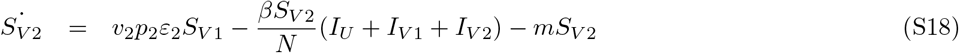

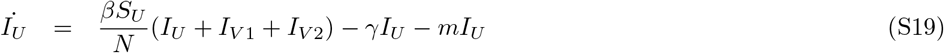

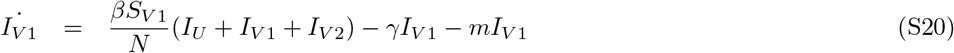

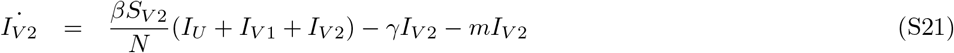

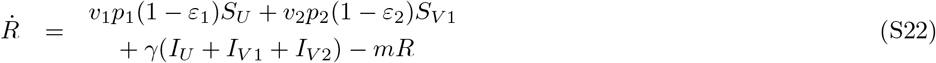

The population is partitioned into seven epidemiological classes: unvaccinated susceptible individuals (*S*_*U*_), unvaccinated infected individuals (*I*_*U*_), individuals who remain susceptible after an unsuccessful immune response to the first dose (*S*_*V* 1_), infected individuals following a single-dose breakthrough (*I*_*V* 1_), individuals who remain susceptible after unsuccessful responses to both doses (*S*_*V* 2_), infected individuals following a double-dose breakthrough (*I*_*V* 2_), and individuals with sterilizing immunity acquired either through infection or successful vaccination (*R*). Individuals enter the population as maternal antibodies wane at rate *b*, which balances removal at the same per capita rate (*b* = *m*). Vaccination occurs at rates *v*_1_ and *v*_2_, corresponding to the average timing of the first and second doses (1*/v*_1_ and 1*/v*_2_ days, respectively). The probabilities of receiving each dose are *p*_1_ and *p*_2_, while *ε*_1_ and *ε*_2_ represent the probabilities of primary vaccine failure following each dose. Note that *ε*_1_*ε*_2_ is the failure rate once the individual receives both doses. Infected individuals in classes *I*_*U*_, *I*_*V* 1_, and *I*_*V* 2_ contribute equally to transmission with coefficient *β*, and individuals in all infected classes recover at per capita rate *γ*.

Our results show that children who have received a single dose (*I*_*V* 1_) experience substantially more infections than fully vaccinated children (*I*_*V* 2_) across all first-dose coverage values (*p*_1_) (Figure S2). This occurs because vaccine failure after the first dose (*ε*_1_ = 0.07) is more common than failure after the full two-dose series (*ε*_1_*ε*_2_ = 0.03), making partially vaccinated children disproportionately represented among infections. These results indicate that reports of primary vaccine failure during outbreaks may often reflect infections in children who have not yet completed the vaccine schedule rather than failures of the full two-dose regimen. Such patterns can therefore give the misleading impression that vaccines that require multiple doses (e.g., MMR) are less effective than they truly are, particularly when breakthrough infections are reported without distinguishing between one-dose and two-dose recipients.ge

### S3 Supplemental figures

**Figure S1:**
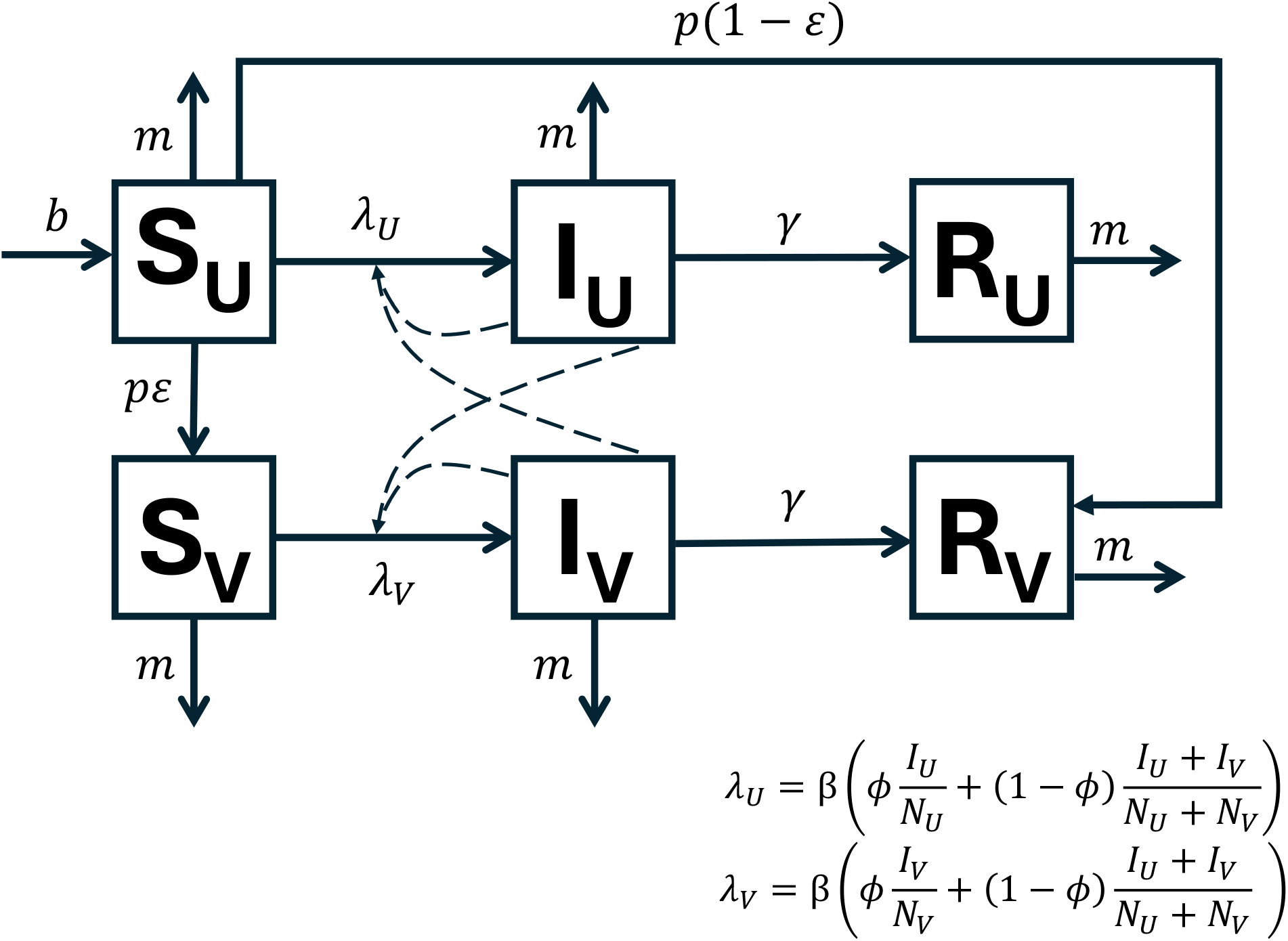
Compartmental diagram of a model with vaccine refusal and breakthrough infections. Boxes indicate states, with capital letters indicating immunity/infection status and subscripts indicating vaccination status. Susceptible individuals are either unvaccinated (*S*_*U*_) or unprotected due to primary vaccine failure (*S*_*V*_). Infected people may either be unvaccinated (*I*_*U*_) or have experienced primary failure (*I*_*V*_). Removed individuals have sterilizing immunity from either infection or successful vaccination. Deaths occur at constant rate *b*, balancing deaths *m*. People elect to get vaccinate with probability *p* and vaccines fail with probability *ε*. New infections arise from contact with infected people regardless of their vaccination status. The disease spreads with force of infection *λ*, a function of transmission coefficient *β*, assortativity *ϕ*, and the current prevalence of infection across the total population of vaccinated and unvaccinated people (*N*_*U*_ and *N*_*V*_ respectively). Recovery occurs at per capita rate *γ*.

**Figure S2:**
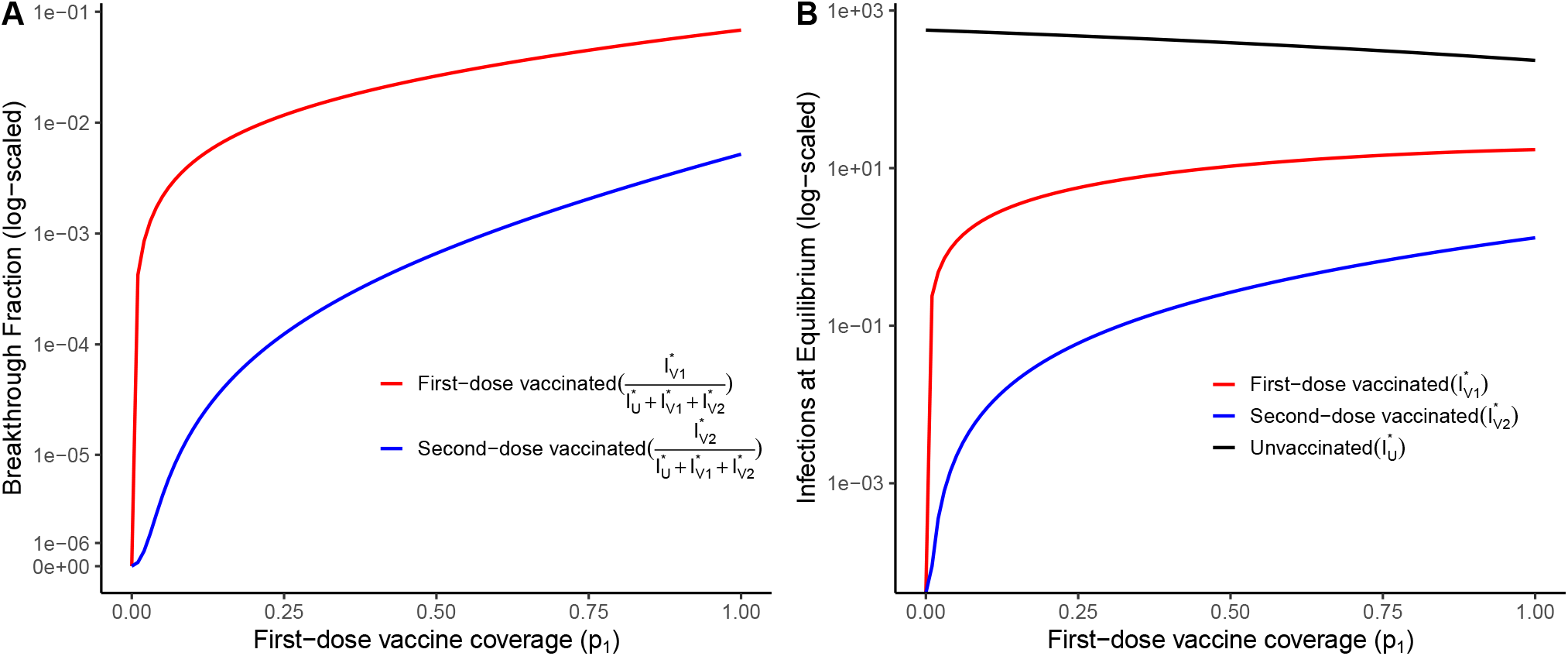
At endemic equilibrium, breakthrough infections disproportionately occur in children with only one dose 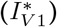 versus the full two-dose regimen 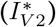 over all theoretical values of first dose coverage (*p*_1_) values. **Left:** As first-dose vaccine coverage (*p*_1_) increases, the fraction of infections occurring in children who have received one dose of the vaccine (red) consistently exceeds the fraction occurring in fully vaccinated children (blue). **Right:** At endemic equilibrium, infections among first-dose recipients remain higher than among second-dose recipients across all *p*_1_ values. Model parameters are consistent with the MMR schedule, post-maternal antibodies wane at six months old: *ε*_1_ = 0.07, *ε*_2_ = 0.43 (two-dose failure *ε*_1_*ε*_2_ = 0.03); 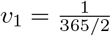(first dose at 1 year old); 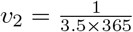 (second dose at 4 years old); 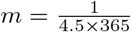 (removal from model once in kindergarten at 5 years old), *β* = 1.515, and *γ* = 0.1, and *p*_2_ = *p*_1_. The population size (*N*) is 100,000.

**Figure S3:**
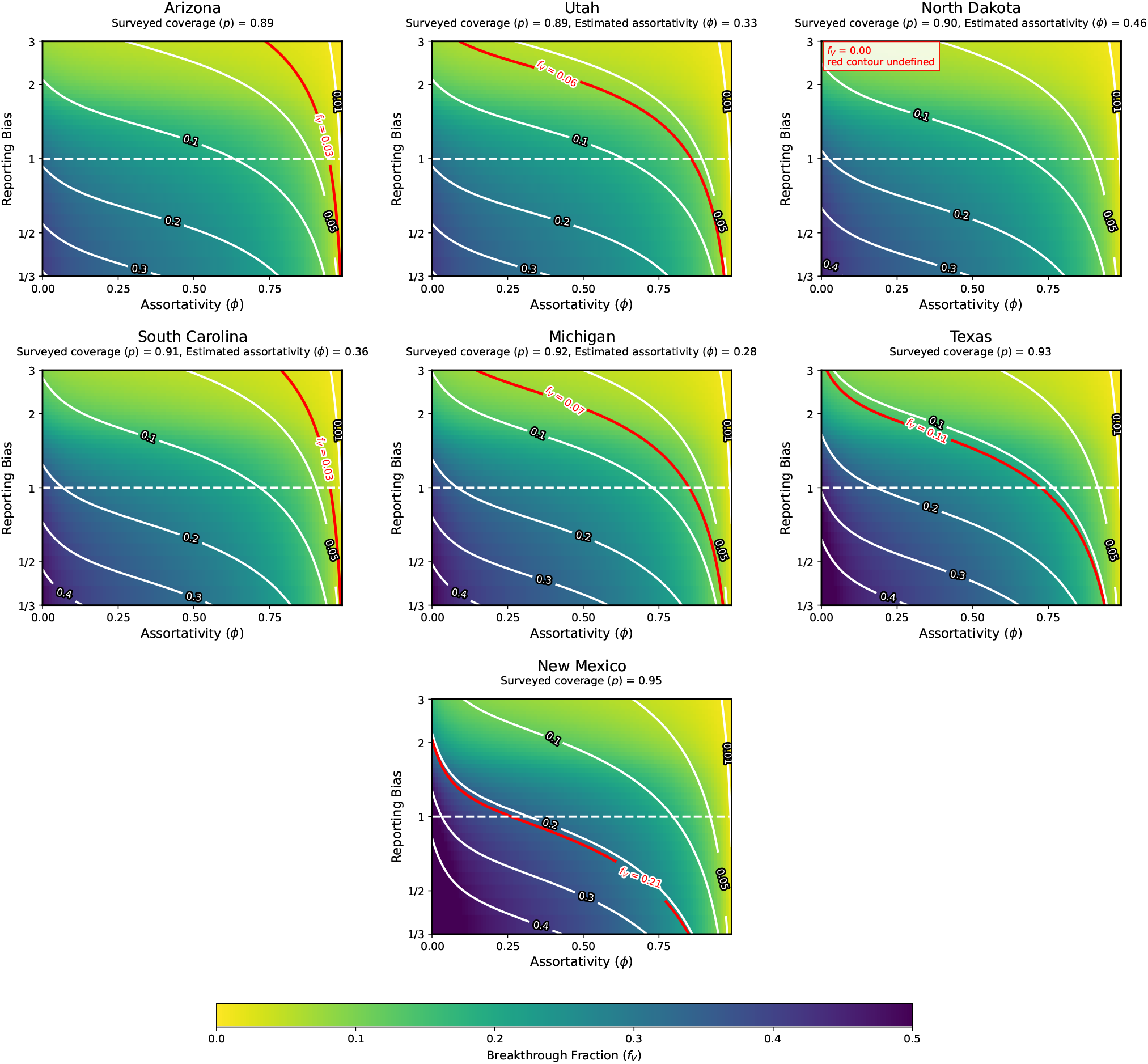
Underreporting of breakthrough infections relative to infections in unvaccinated individuals reduces the observed breakthrough fraction. Heatmaps display the *observed* breakthrough fraction (*f*_*V*_) across different levels of assortativity (*ϕ*, x-axis) and reporting biases (y-axis, log-scaled) depending on surveyed vaccine coverage at the state level (*p*, indicated by facet labels). Reporting bias is defined as the likelihood that a case is detected for an unvaccinated individual relative to a vaccinated individual. The dashed horizontal line corresponds to a reporting bias of one, meaning that case detection is independent of vaccination status. White contour lines indicate the combination of assortativity and reporting bias necessary to return a given breakthrough fraction (indicated by labels; contours provided for breakthrough fraction values of 0.01, 0.05, 0.1, 0.2, 0.3, and 0.4). The red contour lines indicate the observed breakthrough fraction in each state in 2025. Facet subtitles also indicate the mean estimated assortativity for each state based on school-level data where available (see Figure 3 and Figure S6).

**Figure S4:**
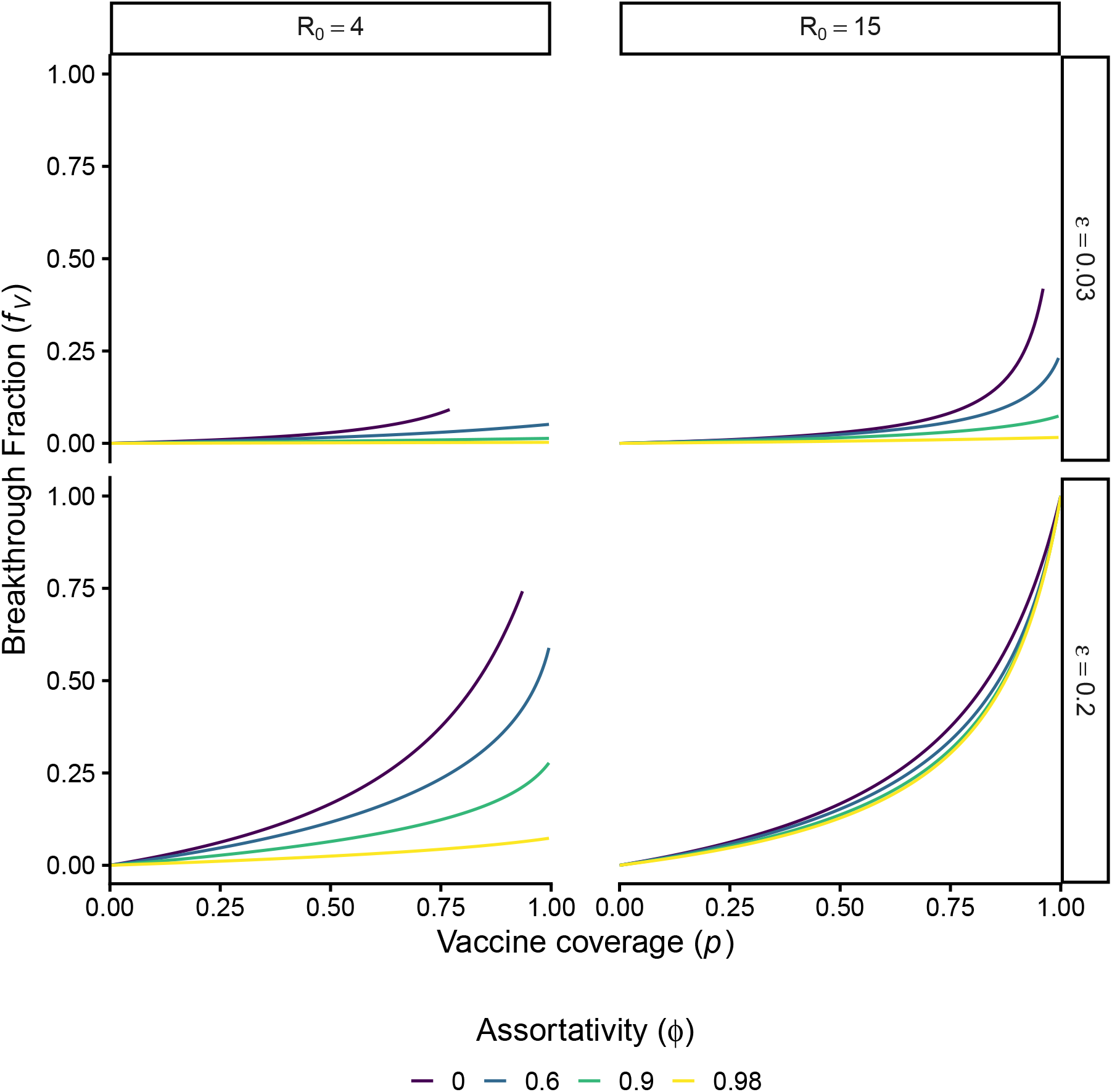
Across different reproduction numbers (*R*_0_) and primary failure rates (*ε*), the fraction of breakthrough infections (*f*_*V*_) increases with vaccine coverage (*p*). Compared to the main analysis (*R*_0_ = 15 and *p* = 0.03), we consider scenarios with a lower reproduction number (*R*_0_ = 4, left column) and a greater primary failure rate (*p* = 0.2, bottom row). Following Figure 2, we show the equilibrium fraction of breakthrough infections across different vaccine coverages (*p*). Colors indicate assortativity (*ϕ*), ranging from uniform mixing (*ϕ* = 0) to high assortativity (*ϕ*=0.98) in a population of one million people.

**Figure S5:**
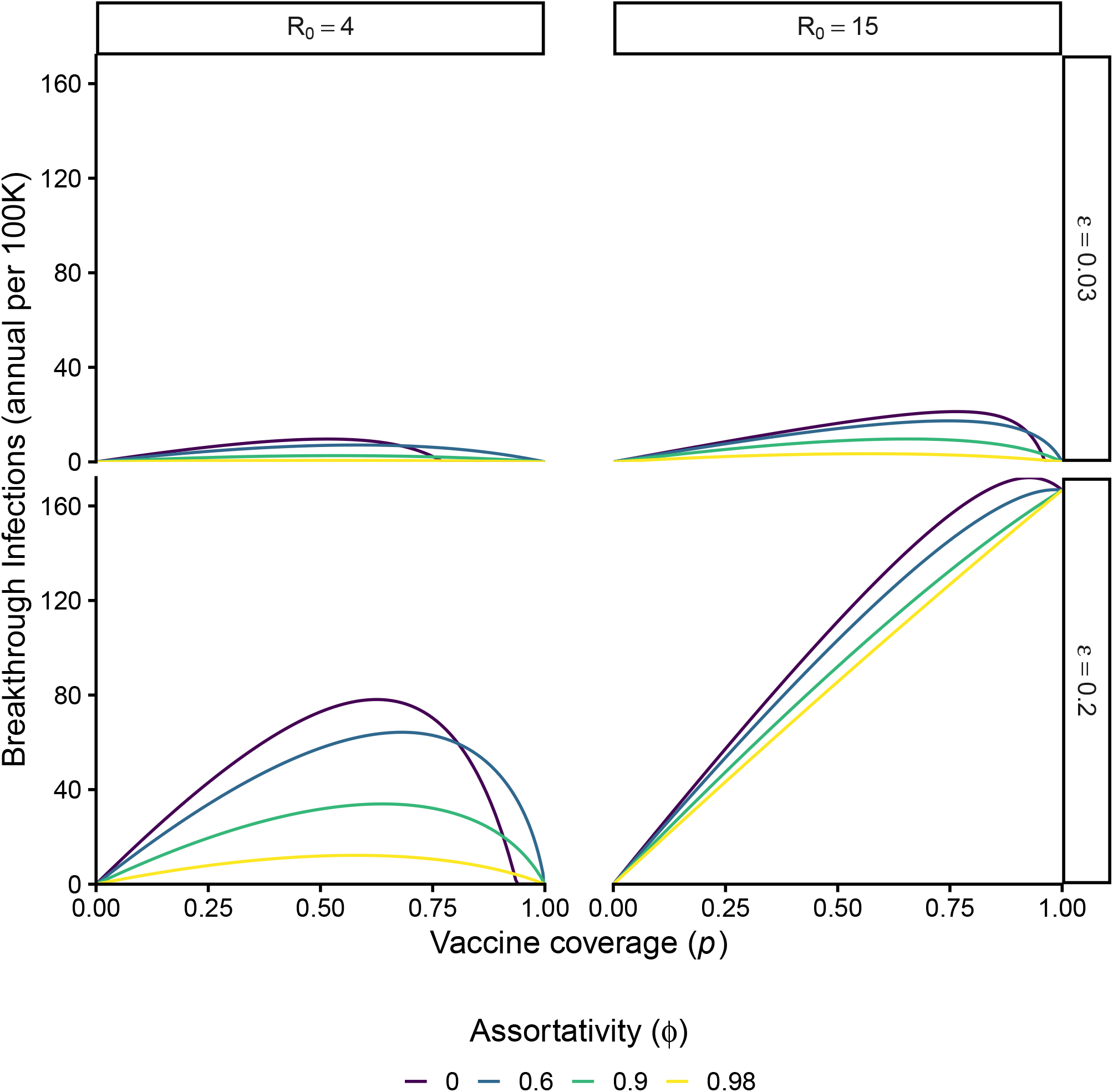
Across different reproduction numbers (*R*_0_) and primary failure rates (*ε*), breakthrough infections (*I*_*V*_) peak at intermediate vaccine coverage (*p*). Compared to the main analysis (*R*_0_ = 15 and *p* = 0.03), we consider scenarios with a lower reproduction number (*R*_0_ = 4, left column) and a greater primary failure rate (*p* = 0.2, bottom row). Following Figure 4, we show the equilibrium number of breakthrough infections across different vaccine coverages (*p*). Colors indicate assortativity (*ϕ*), ranging from uniform mixing (*ϕ* = 0) to high assortativity (*ϕ*=0.98) in a population of one million people.

**Figure S6:**
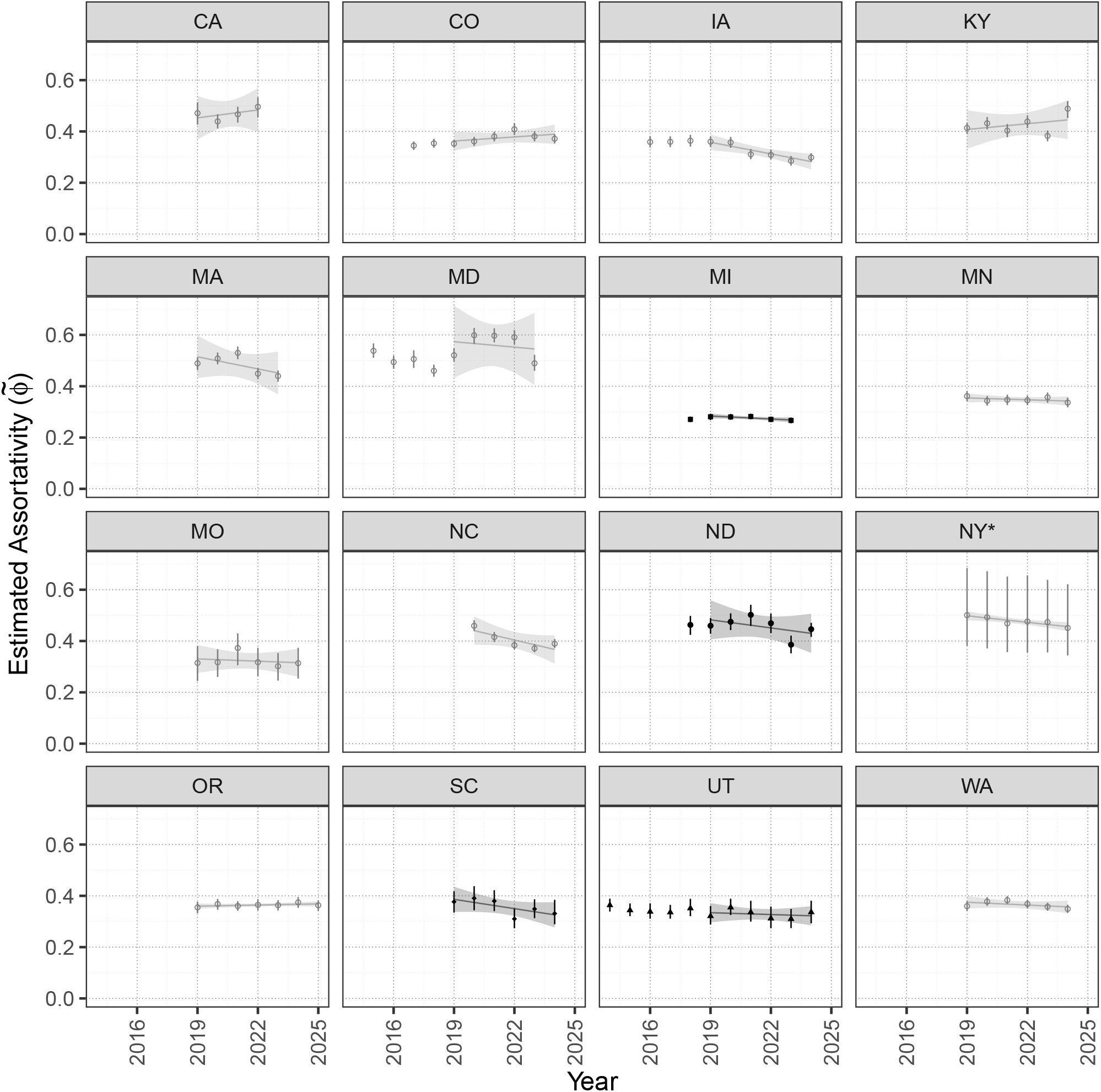
Estimated assortativity is moderate across states and consistent across years since 2019. Points indicate the population-weighted estimated assortativity 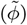 annually and across sixteen states (indicated by facet labels). Years correspond to the start of each school year. Vertical bars correspond to a 95% confidence interval across 1,000 bootstrapped estimates with replacement. The lighter lines indicate the linear trend in assortativity over time since 2019 – moderate to insignificant in all cases. The four states in darker grey (ND, MI, SC, and UT) also had measles epidemics in 2025 and were included in the analysis of breakthrough fractions (see Figure 1 and Figure 2). *New York excludes New York City. See Table S5 for information on data sources and limitations.

**Figure S7:**
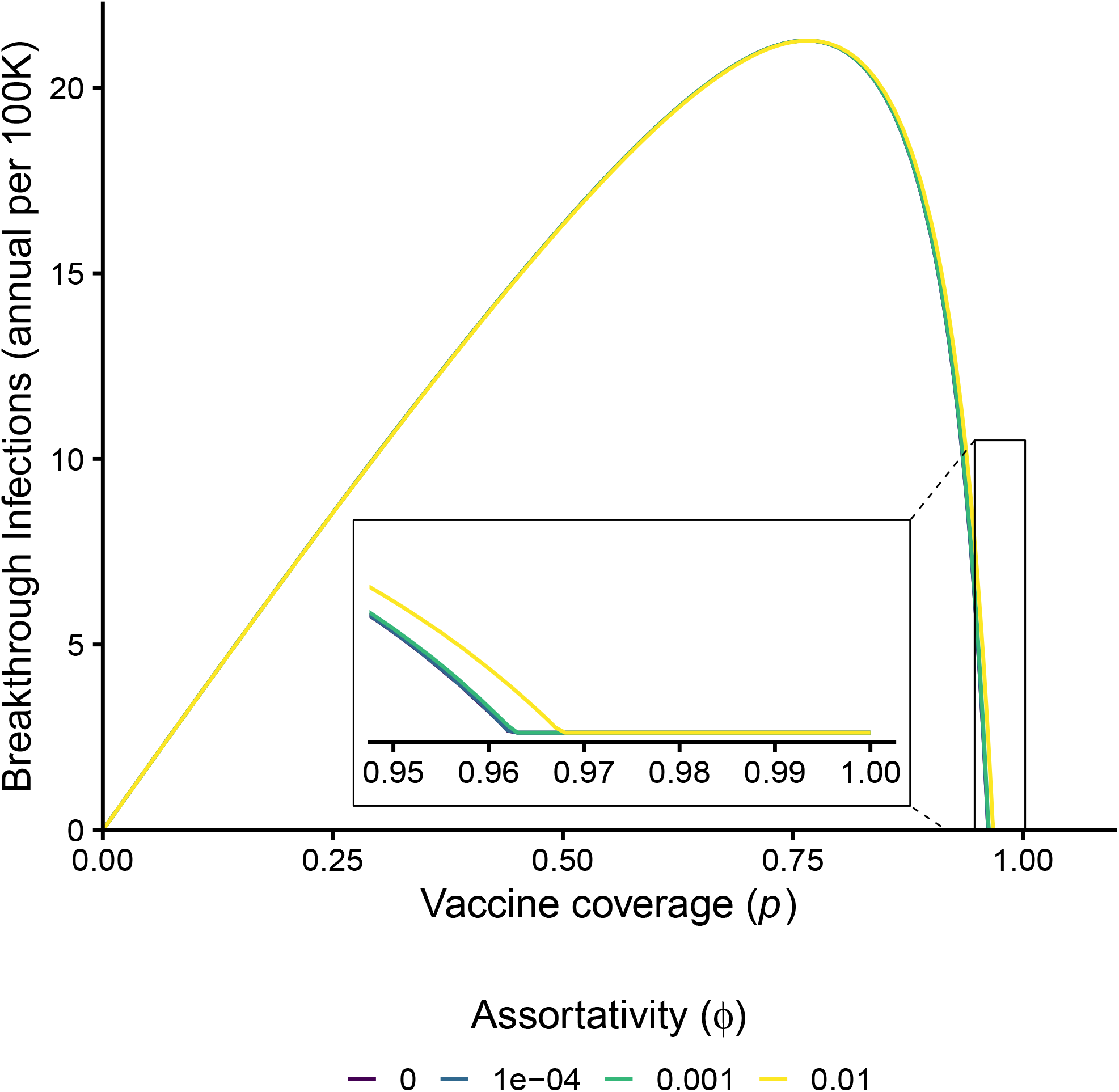
Even mild assortativity necessitates extreme high vaccine coverage to achieve herd immunity. As in Figure 4, lines indicate annual new breakthrough cases per 100K people depending on vaccine coverage (*p*) and assortativity (*ϕ*, colors). Assortativity values now vary by orders of magnitude from 10^−4^ to 0.1. The inset zooms focuses on coverage above 0.95 to emphasize the herd immunity threshold. Parameters are consistent with measles and the MMR vaccine (i.e., *R*_0_ = 15 and failure rate *ϵ* = 0.03, see Table S1).

### S4 Supplemental tables

**Table S1:** Parameters for model described by Equation S1.

| Parameter | Symbol | Value |
| --- | --- | --- |
| Total population size | $N_T$ | $10^7$ |
| Per capita recovery rate | $\gamma$ | $0.1 \text{ days}^{-1}$ |
| Per capita death rate | $m$ | $1/80 \text{ years}^{-1}$ |
| Basic reproduction number | $\mathcal{R}_0$ | $\{4, 15\}$ |
| Birth rate | $b$ | $mN_T$ |
| Transmission coefficient | $\beta$ | $\mathcal{R}_0(\gamma + m)$ |
| Vaccine coverage | $p$ | $[0, 1]$ |
| Vaccine failure probability | $\varepsilon$ | $\{0.03, 0.2\}$ |
| Assortativity | $\phi$ | $\{0, 0.3, 0.6, 0.9, 0.98\}$ |

**Table S2:** Initial conditions for numerical integration of Equation S1.

| State variable | Symbol | Value |
| --- | --- | --- |
| Unvaccinated susceptible | $S_U(t=0)$ | $(1-p)(N_T - 100)$ |
| Vaccinated susceptible | $S_V(t=0)$ | $p(N_T - 100)$ |
| Unvaccinated infectious | $I_U(t=0)$ | $100(1-p)$ |
| Vaccinated infectious | $I_V(t=0)$ | $100p$ |
| Unvaccinated recovered | $R_U(t=0)$ | 0 |
| Vaccinated recovered | $R_V(t=0)$ | 0 |

**Table S3:**
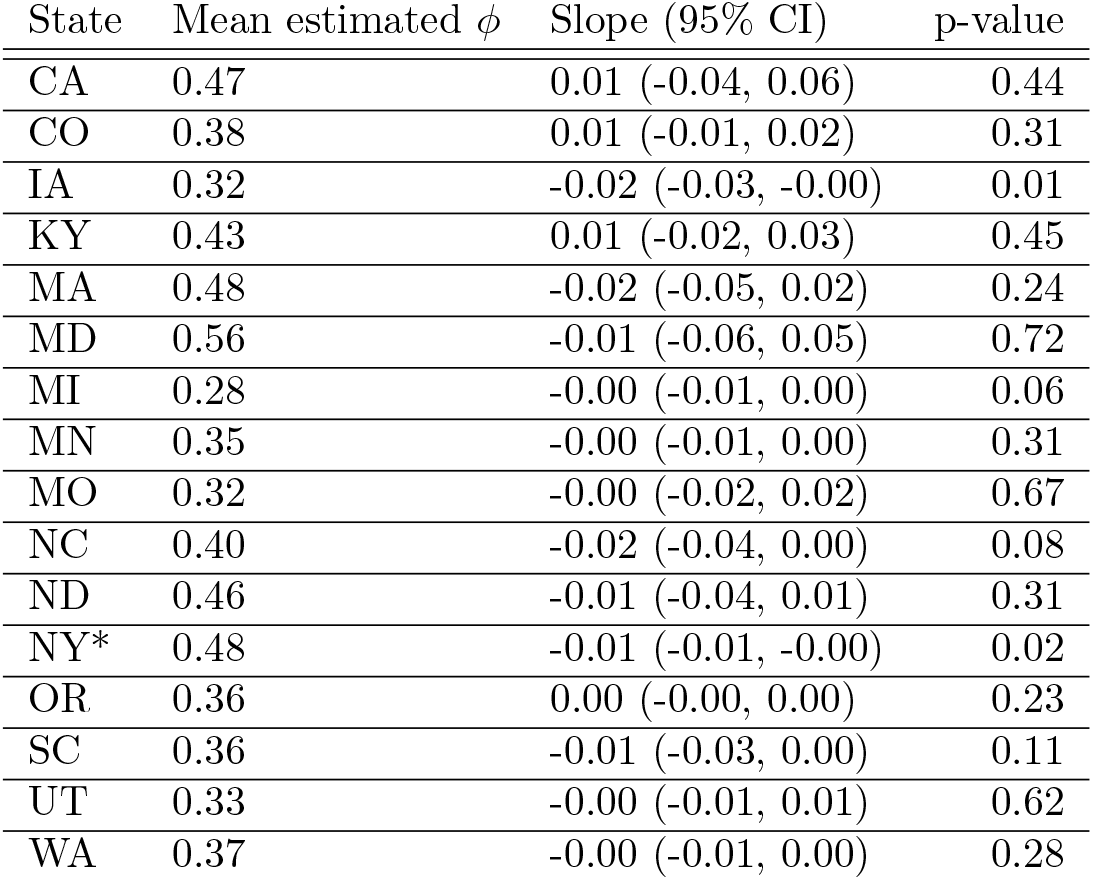
No significant trend in assortativity since 2019. We estimated assortativity (*ϕ*) based on school-level MMR coverage in kindergarteners. Columns indicate the state postal abbreviation, the mean estimate *ϕ* since 2019, the estimated slope of *ϕ* over time since 2019, and the p-value of the two-sided t-test for the slope (i.e., Pr(—t— *>* 0)). * New York excludes schools in New York City.

| State | Mean estimated $\phi$ | Slope (95% CI) | p-value |
| --- | --- | --- | --- |
| CA | 0.47 | 0.01 (-0.04, 0.06) | 0.44 |
| CO | 0.38 | 0.01 (-0.01, 0.02) | 0.31 |
| IA | 0.32 | -0.02 (-0.03, -0.00) | 0.01 |
| KY | 0.43 | 0.01 (-0.02, 0.03) | 0.45 |
| MA | 0.48 | -0.02 (-0.05, 0.02) | 0.24 |
| MD | 0.56 | -0.01 (-0.06, 0.05) | 0.72 |
| MI | 0.28 | -0.00 (-0.01, 0.00) | 0.06 |
| MN | 0.35 | -0.00 (-0.01, 0.00) | 0.31 |
| MO | 0.32 | -0.00 (-0.02, 0.02) | 0.67 |
| NC | 0.40 | -0.02 (-0.04, 0.00) | 0.08 |
| ND | 0.46 | -0.01 (-0.04, 0.01) | 0.31 |
| NY* | 0.48 | -0.01 (-0.01, -0.00) | 0.02 |
| OR | 0.36 | 0.00 (-0.00, 0.00) | 0.23 |
| SC | 0.36 | -0.01 (-0.03, 0.00) | 0.11 |
| UT | 0.33 | -0.00 (-0.01, 0.01) | 0.62 |
| WA | 0.37 | -0.00 (-0.01, 0.00) | 0.28 |

**Table S4:**
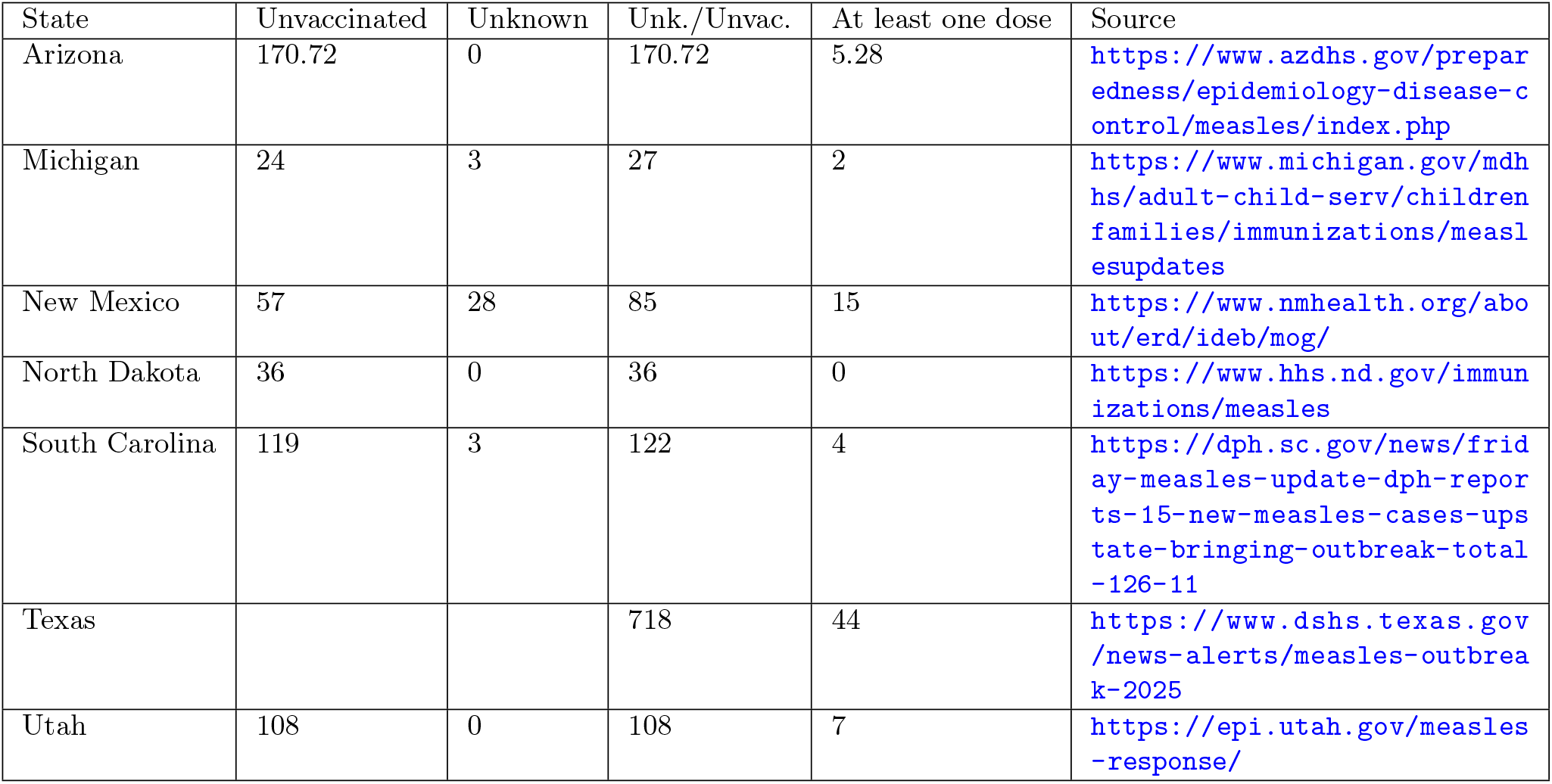
State-level data on measles infections by December 15, 2025 by vaccination status. Columns provide the number of infections by vaccination status: unvaccinated, unknown, unknown or unvaccinated (abbreviated as Unk./Unvac.; equivalent to the prior two categories combined), and at least one dose, as well as the source for the data. Texas did not distinguish between unvaccinated people and people with unknown vaccination status. Values for Michigan distinguishing between unvaccinated and unknown status were provided via correspondence with the Michigan Department of Health and Human Services. Values for Arizona are nonintegers; reported percentages were multiplied by total reported cases to estimate counts.

**Table S5:**
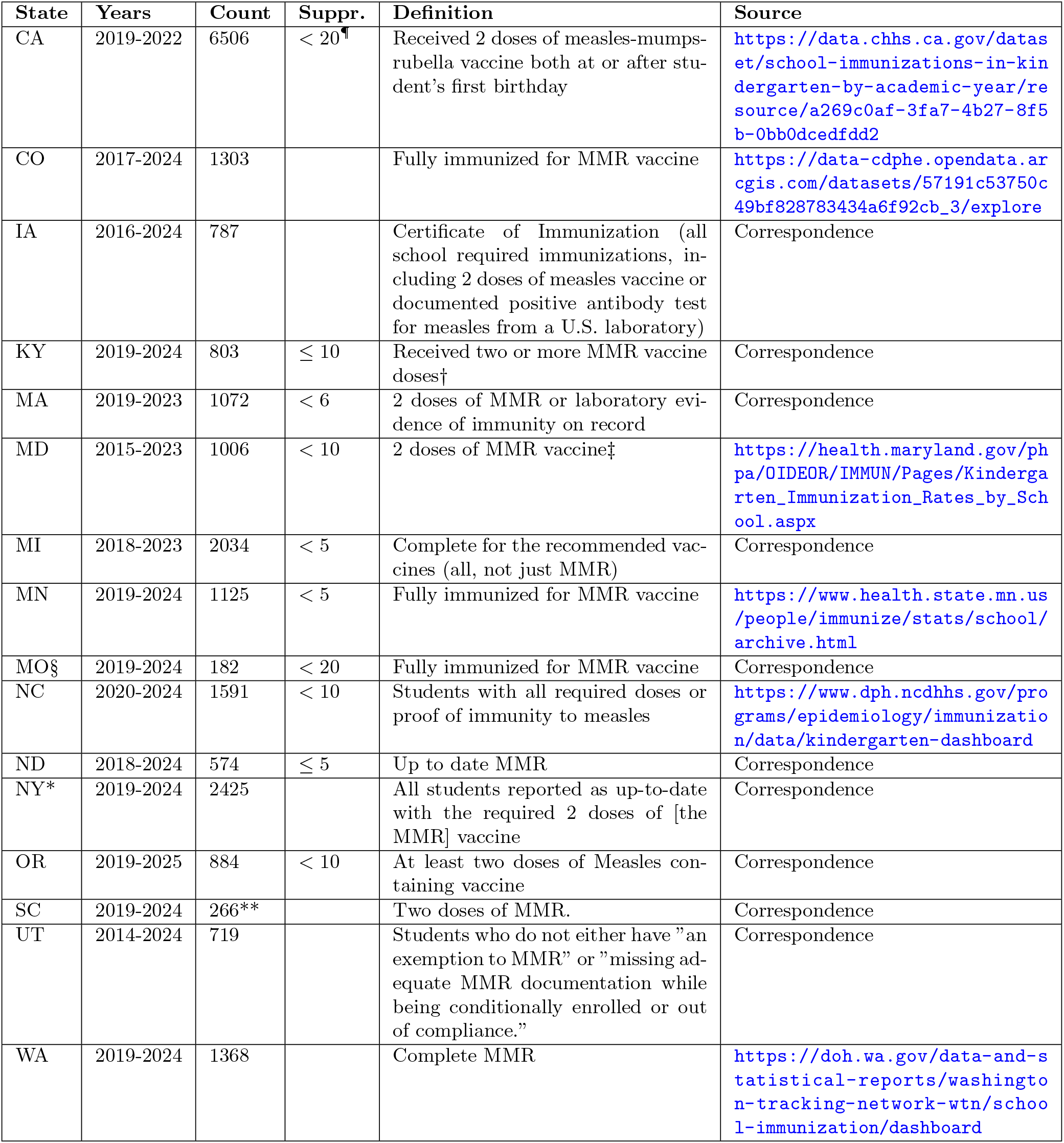
School-level vaccination data information. From left to right, columns indicate the state abbreviation, the years of data available, the average number of schools (or zip codes, see §) per year, the criteria for suppression (abbreviated as Suppr., meaning removal of small units to preserve anonymity), the definition of coverage, and the source of data. ^¶^: Binning is applied when a small percentage of students are vaccinated or unvaccinated (for example,”in schools with 20-49 enrollees, values of 95% or higher are listed as *>* 95%, and values of 5% or less are listed as *<* 5%”). ^*†*^: Starting in 2024-2025, vaccine rates are now based on age- and interval-specific criteria rather than (as in previous years) simply counting the number of doses received regardless of when they were administered. ‡: “Children reporting a medical or religious exemption are not included in the coverage rate calculations for each vaccine.” To correct for this, we assume that students with exemptions are unvaccinated and recalculate coverage accordingly: *p* = (1 − %*EXEMPT*) ^*^ *REPORTED*.*COV ERAGE* §: Zipcode-level data. “Zip codes are based on the location of the reporting school district/school to the Missouri Department of Elementary & Secondary Education (DESE).” *: New York State excluding New York City **: Sample of schools. “This data comes from the Department of Public Health’s 5K audits, which are done annually by our immunization compliance staff. The 5K audits are conducted on a sample size of the kindergarten population. The sample size is a minimum of 385 5K students per county (schools with 5K enrollment are randomly selected until the minimum sample size is reached). In the smaller counties with less than 385 total 5K students, all schools are selected for audit.”

## Notes

### Competing Interest Statement

The authors have declared no competing interest.

### Summary of Updates

Supplemental analysis examining the potential effects of biases in case detection depending on vaccination status (Fig S3)

## References

C. L. Althaus and M. Salathé. Measles vaccination coverage and cases among vaccinated persons. Emerg. Infect. Dis., 21(8):1480–1481, Aug. 2015.

R. Anderson and R. May. Infectious diseases of humans: Dynamics and control. Oxford University Press, 1991.

K. L. Andrejko, J. R. Head, J. A. Lewnard, and J. V. Remais. Longitudinal social contacts among school-aged children during the COVID-19 pandemic: the bay area contacts among kids (BACK) study. BMC Infect. Dis., 22(1):242, Mar. 2022.

Y. Arima and K. Oishi. Letter to the editor: Measles cases among fully vaccinated persons. Euro Surveill., 23 (34), Aug. 2018.

R. A. Bednarczyk and M. E. Sundaram. The continued risk of measles outbreaks in the United States resulting from suboptimal vaccination coverage. Public Health Rep., page 333549241306608, Jan. 2025.

D. Bhatia, N. Crowcroft, S. Antoni, M. C. Danovaro-Holliday, A. S. Bose, A. Minta, B. Masresha, and M. J. Ferrari. Prediction of subnational-level vaccination coverage estimates using routine surveillance data and survey data. Vaccine, 60(127277):127277, July 2025.

S. Busenberg and C. Castillo-Chavez. A general solution of the problem of mixing of subpopulations and its application to risk- and age-structured epidemic models for the spread of AIDS. Math. Med. Biol., 8(1):1–29, 1991.

R. M. Carpiano, T. Callaghan, R. DiResta, N. T. Brewer, C. Clinton, A. P. Galvani, R. Lakshmanan, W. E. Parmet, S. B. Omer, A. M. Buttenheim, R. M. Benjamin, A. Caplan, J. A. Elharake, L. C. Flowers, Y. A. Maldonado, M. M. Mello, D. J. Opel, D. A. Salmon, J. L. Schwartz, J. M. Sharfstein, and P. J. Hotez. Confronting the evolution and expansion of anti-vaccine activism in the USA in the COVID-19 era. Lancet, 401(10380):967–970, Mar. 2023.

A. Cassini, L. Cobuccio, E. Glampedakis, P. Cherpillod, P. A. Crisinel, F.-J. Pérez-Rodríguez, M. Attinger, D. Bachelin, M. N. Tessemo, M. Maeusezahl, C. Gardiol, and K. Boubaker. Adapting response to a measles outbreak in a context of high vaccination and breakthrough cases: an example from Vaud, Switzerland, January to March 2024. Euro Surveill., 29(22), May 2024.

Centers for Disease Control and Prevention. Measles vaccination, 2025a. URL https://www.cdc.gov/measles/vaccines/index.html. Accessed: 2025-11-12.

Centers for Disease Control and Prevention. Estimated vaccination coverage for MMR vaccine, not up-to-date, and exempt from one or more vaccines among children enrolled in kindergarten, by jurisdiction — United States, 2024–25 school year, 2025b. URL https://www.cdc.gov/schoolvaxview/media/files/2025/07/SchoolVaxView_Weighted_Counts_Table_20250728.xlsx. Accessed: 2025-12-16.

S. Chen and A. I. Bento. Multiscale modelling reveals accelerating community outbreak risks of measles in the united states. Jan. 2026.

J. D. Cherry and M. Zahn. Clinical characteristics of measles in previously vaccinated and unvaccinated patients in California. Clin. Infect. Dis., 67(9):1315–1319, Oct. 2018.

L. Cunniff, E. Alyanak, A. Fix, M. Novak, M. Peterson, K. Mevis, A. L. Eiden, and A. Bhatti. The impact of the COVID-19 pandemic on vaccination uptake in the united states and strategies to recover and improve vaccination rates: A review. Hum. Vaccin. Immunother., 19(2):2246502, Aug. 2023.

G. De Serres, N. Boulianne, F. Meyer, and B. J. Ward. Measles vaccine efficacy during an outbreak in a highly vaccinated population: incremental increase in protection with age at vaccination up to 18 months. Epidemiol. Infect., 115(2):315–323, Oct. 1995.

G. De Serres, N. Boulianne, F. Defay, N. Brousseau, M. Benoît, S. Lacoursiére, F. Guillemette, J. Soto, M. Ouakki, B. J. Ward, and D. M. Skowronski. Higher risk of measles when the first dose of a 2-dose schedule of measles vaccine is given at 12-14 months versus 15 months of age. Clin. Infect. Dis., 55(3):394–402, Aug. 2012.

S. B. Dolan, E. Carnahan, J. C. Shearer, E. N. Beylerian, J. Thompson, S. S. Gilbert, L. Werner, and T. K. Ryman. Redefining vaccination coverage and timeliness measures using electronic immunization registry data in low- and middle-income countries. Vaccine, 37(13):1859–1867, Mar. 2019.

E. Dong, A. Nearchou, Y. Okura, S. Saiyed, and L. M. Gardner. MMR vaccination coverage in the U.S. before and after the COVID-19 pandemic: A modelling study. Feb. 2025a.

E. Dong, S. Saiyed, A. Nearchou, Y. Okura, and L. M. Gardner. Trends in county-level MMR vaccination coverage in children in the United States. JAMA, 334(8):730–732, Aug. 2025b.

J. Dushoff and S. Levin. The effects of population heterogeneity on disease invasion. Math. Biosci., 128(1-2): 25–40, July 1995.

I. Evans, S. Jury, A. Morrison, E. Best, V. King, and E. Reynolds. Onward transmission of measles virus among vaccinated cases in a large community outbreak in Auckland, New Zealand, 2019. Vaccine, 42(23):126257, Oct. 2024.

M. Fattah, L. A. Stoffel, K. M. Bubar, S. J. Bents, Y. Maldonado, P. J. Hotez, M. V. Kiang, and N. C. Lo. Trends in county-level childhood vaccination exemptions in the us. JAMA, Jan. 2026. ISSN 0098-7484. doi: 10.1001/jama.2025.24407. URL http://dx.doi.org/10.1001/jama.2025.24407.

M. C. Fitzpatrick, C. R. Wells, A. Pandey, L. Ayaz, P. J. Hotez, S. M. Moghadas, and A. P. Galvani. School-level gaps in MMR coverage as the fuel for measles outbreaks. Ann. Intern. Med., (ANNALS-25-01611), Oct. 2025.

P. A. Gastañaduy, S. Funk, B. A. Lopman, P. A. Rota, M. Gambhir, B. Grenfell, and P. Paul. Factors associated with measles transmission in the United States during the postelimination era. JAMA Pediatr., 174(1):56–62, Jan. 2020.

U. Heininger, N. S. Bachtiar, P. Bahri, A. Dana, A. Dodoo, J. Gidudu, and E. M. D. Santos. The concept of vaccination failure. Vaccine, 30(7):1265–1268, Feb. 2012.

D. R. Hijano, W. A. Orenstein, and C. R. Oliveira. Measles resurgence and the fragility of herd immunity: Implications for pediatric infectious disease practice. J. Pediatric Infect. Dis. Soc., 14(11), Nov. 2025.

T. Hiraoka, A. K. Rizi, M. Kivelä, and J. Saramäki. Herd immunity and epidemic size in networks with vaccination homophily. Phys. Rev. E., 105(5):L052301, May 2022.

A. Kata. A postmodern Pandora’s box: anti-vaccination misinformation on the internet. Vaccine, 28(7):1709–1716, Feb. 2010.

M. V. Kiang, K. M. Bubar, Y. Maldonado, P. J. Hotez, and N. C. Lo. Modeling reemergence of vaccine-eliminated infectious diseases under declining vaccination in the US. JAMA, 333(24):2176–2187, June 2025.

C. W. LeBaron, J. Beeler, B. J. Sullivan, B. Forghani, D. Bi, C. Beck, S. Audet, and P. Gargiullo. Persistence of measles antibodies after 2 doses of measles vaccine in a postelimination environment. Archives of pediatrics & adolescent medicine, 161(3):294–301, 2007.

D. I. Lee, A. Nande, T. L. Anderson, M. Z. Levy, and A. L. Hill. Vaccine failure mode determines population-level impact of vaccination campaigns during epidemics. J. R. Soc. Interface, 22(223):20240689, Feb. 2025.

J. Leung, N. A. Munir, A. D. Mathis, T. D. Filardo, P. A. Rota, D. E. Sugerman, S. B. Sowers, S. Mercader, S. N. Crooke, and P. A. Gastañaduy. The effects of vaccination status and age on clinical characteristics and severity of measles cases in the United States in the postelimination era, 2001-2022. Clin. Infect. Dis., 80(3): 663–672, Mar. 2025.

F. A. Lievano, M. J. Papania, R. F. Helfand, R. Harpaz, L. Walls, R. S. Katz, I. Williams, Y. S. Villamarzo, P. A. Rota, and W. J. Bellini. Lack of evidence of measles virus shedding in people with inapparent measles virus infections. J. Infect. Dis., 189 Suppl 1(Supplement 1):S165–70, May 2004.

N. B. Masters, M. C. Eisenberg, P. L. Delamater, M. Kay, M. L. Boulton, and J. Zelner. Fine-scale spatial clustering of measles nonvaccination that increases outbreak potential is obscured by aggregated reporting data. Proc. Natl. Acad. Sci. U. S. A., 117(45):28506–28514, Nov. 2020.

H. Q. McLean, A. P. Fiebelkorn, J. L. Temte, and G. S. Wallace. Prevention of measles, rubella, congenital rubella syndrome, and mumps, 2013: summary recommendations of the advisory committee on immunization practices (acip). MMWR. Recommendations and reports: Morbidity and mortality weekly report. Recommendations and reports, 62(RR-04):1–34, 2013.

J. D. Munday, K. E. Atkins, D. Klinkenberg, M. Meurs, E. Fleur, S. J. Hahné, J. Wallinga, and A. Jan van Hoek. Estimating the risk and spatial spread of measles in populations with high MMR uptake: Using school-household networks to understand the 2013 to 2014 outbreak in the Netherlands. PLoS Med., 21(10):e1004466, Oct. 2024.

K. N. Nelson, A. J. Siegler, P. S. Sullivan, H. Bradley, E. Hall, N. Luisi, P. Hipp-Ramsey, T. Sanchez, K. Shioda, and B. A. Lopman. Nationally representative social contact patterns among U.S. adults, August 2020-April 2021. Sept. 2021.

New Mexico Department of Health. 2025 measles outbreak guidance. https://www.nmhealth.org/about/erd/ideb/mog/, 2025. Accessed: 2025-12-08.

S. R. Newcomer, J. Graham, K. Irish, R. E. Freeman, C. S. Leary, B. K. Wehner, and M. F. Daley. Identification of spatial clusters of undervaccination patterns among children aged <24 months using immunization information system data, Montana, 2015-2019. Public Health Rep., 139(3):360–368, May 2024.

W. A. Orenstein, R. H. Bernier, T. J. Dondero, A. R. Hinman, J. S. Marks, K. J. Bart, and B. Sirotkin. Field evaluation of vaccine efficacy. Bull. World Health Organ., 63(6):1055–1068, 1985.

Pan American Health Organization. Paho calls for regional action as the Americas lose measles elimination status, 2025. URL https://www.paho.org/en/news/10-11-2025-paho-calls-regional-action-americas-lose-measles-elimination-status. Accessed: 2025-11-12.

S. W. Park, D. M. Cornforth, J. Dushoff, and J. S. Weitz. The time scale of asymptomatic transmission affects estimates of epidemic potential in the COVID-19 outbreak. Epidemics, 31:100392, 2020.

M. Paunio, K. Hedman, I. Davidkin, and H. Peltola. IgG avidity to distinguish secondary from primary measles vaccination failures: prospects for a more effective global measles elimination strategy. Expert Opinion on Pharmacotherapy, 4(8):1215–1225, 2003.

I. R. Pedersen, C. H. Mordhorst, G. Glikmann, and H. von Magnus. Subclinical measles infection in vaccinated seropositive individuals in arctic greenland. Vaccine, 7(4):345–348, Aug. 1989.

I. Pedroza-Meza, M. A. Acuña-Zegarra, and J. X. Velasco-Hernández. Modeling vaccine failures and behavioral change: Effects on disease transmission dynamics and thresholds. Math. Biosci., (109619):109619, Jan. 2026.

G. A. Poland. Failure to reach the goal of measles elimination. Arch. Intern. Med., 154(16):1815, Aug. 1994.

G. A. Poland and R. M. Jacobson. The re-emergence of measles in developed countries: time to develop the next-generation measles vaccines? Vaccine, 30(2):103–104, Jan. 2012.

Posit team. RStudio: Integrated Development Environment for R. Posit Software, PBC, Boston, MA, 2025. URL http://www.posit.co/.

R Core Team. R: A Language and Environment for Statistical Computing. R Foundation for Statistical Computing, Vienna, Austria, 2021. URL https://www.R-project.org/.

E. Rahimi, E. Ghaderi, E. Mostafavi, and M. Karami. The quality of measles outbreak investigation report, how can it bridge the gap and help to fulfill the goal of measles elimination? BMC Infect. Dis., 25(1):496, Apr. 2025.

A. Robert, A. M. Suffel, and A. J. Kucharski. Long-term waning of vaccine-induced immunity to measles in england: a mathematical modelling study. Lancet Public Health, 9(10):e766–e775, Oct. 2024.

D. A. Salmon, P. J. Smith, A. M. Navar, W. K. Y. Pan, S. B. Omer, J. A. Singleton, and N. A. Halsey. Measuring immunization coverage among preschool children: past, present, and future opportunities. Epidemiol. Rev., 28(1):27–40, June 2006.

N. Sundell, L. Dotevall, M. Sansone, M. Andersson, M. Lindh, T. Wahlberg, T. Tyrberg, J. Westin, J.-Å. Liljeqvist, T. Bergström, M. Studahl, and L.-M. Andersson. Measles outbreak in Gothenburg urban area, Sweden, 2017 to 2018: low viral load in breakthrough infections. Euro Surveill., 24(17), Apr. 2019.

P. A. Sutcliffe and E. Rea. Outbreak of measles in a highly vaccinated secondary school population. CMAJ, 155 (10):1407–1413, Nov. 1996.

J. C. Taube, Z. Susswein, V. Colizza, and S. Bansal. Characterising non-household contact patterns relevant to respiratory transmission in the USA: analysis of a cross-sectional survey. Lancet Digit. Health, 7(8):100888, Aug. 2025.

The MathWorks, Inc. MATLAB version R2025b, 2025. URL https://www.mathworks.com.

S. A. Truelove, M. Graham, W. J. Moss, C. J. E. Metcalf, M. J. Ferrari, and J. Lessler. Characterizing the impact of spatial clustering of susceptibility for measles elimination. Vaccine, 37(5):732–741, Jan. 2019.

A. Uzicanin and L. Zimmerman. Field effectiveness of live attenuated measles-containing vaccines: a review of published literature. J. Infect. Dis., 204 Suppl 1(suppl 1):S133–48, July 2011.

C. R. Vitek, M. Aduddell, M. J. Brinton, R. E. Hoffman, and S. C. Redd. Increased protections during a measles outbreak of children previously vaccinated with a second dose of measles-mumps-rubella vaccine. Pediatr. Infect. Dis. J., 18(7):620–623, July 1999.

S. E. Wilson, A. Bunko, S. Johnson, J. Murray, Y. Wang, S. L. Deeks, N. S. Crowcroft, L. Friedman, L. C. Loh, M. MacLeod, C. Taylor, and Y. Li. The geographic distribution of un-immunized children in Ontario, Canada: Hotspot detection using Bayesian spatial analysis. Vaccine, 39(8):1349–1357, Feb. 2021.

L. Yang, B. T. Grenfell, and M. J. Mina. Waning immunity and re-emergence of measles and mumps in the vaccine era. Curr. Opin. Virol., 40:48–54, Feb. 2020.

L. F. Yeung, P. Lurie, G. Dayan, E. Eduardo, P. H. Britz, S. B. Redd, M. J. Papania, and J. F. Seward. A limited measles outbreak in a highly vaccinated US boarding school. Pediatrics, 116(6):1287–1291, Dec. 2005.

E. G. Zhou, J. S. Brownstein, and B. Rader. Assessing MMR vaccination coverage gaps in US children with digital participatory surveillance. Nature Health, 1(1):138–144, Jan. 2026. ISSN 3005-0693. doi: 10.1038/s44360-025-00031-8. URL http://dx.doi.org/10.1038/s44360-025-00031-8.

